# Environmental and genetic predictors of human cardiovascular ageing

**DOI:** 10.1101/2022.10.03.22280042

**Authors:** Mit Shah, Marco H. de A. Inácio, Chang Lu, Pierre-Raphaël Schiratti, Sean L. Zheng, Adam Clement, Wenjia Bai, Andrew P. King, James S. Ware, Martin R. Wilkins, Johanna Mielke, Eren Elci, Ivan Kryukov, Kathryn A. McGurk, Christian Bender, Daniel F. Freitag, Declan P. O’Regan

## Abstract

Cardiovascular ageing is a process that begins early in life and leads to a progressive change in structure and decline in function due to accumulated damage across diverse cell types, tissues and organs contributing to multi-morbidity. Damaging biophysical, metabolic and immunological factors exceed endogenous repair mechanisms resulting in a pro-fibrotic state, cellular senescence and end-organ damage, however the genetic architecture of cardiovascular ageing is not known. Here we used machine learning approaches to quantify cardiovascular age from image-derived traits of vascular function, cardiac motion and myocardial fibrosis, as well as conduction traits from electrocardiograms, in 39,559 participants of UK Biobank. Cardiovascular ageing was found to be significantly associated with common or rare variants in genes regulating sarcomere homeostasis, myocardial immunomodulation, and tissue responses to biophysical stress. Ageing is accelerated by cardiometabolic risk factors and we also identified prescribed medications that were potential modifiers of ageing. Through large-scale modelling of ageing across multiple traits our results reveal insights into the mechanisms driving premature cardiovascular ageing and reveal potential molecular targets to attenuate age-related processes.

## Introduction

Cardiovascular disease (CVD) is the leading cause of death globally, and ageing is a primary risk factor for its development and progression.^1,2^ Cardiovascular ageing is a process that begins in early life and occurs at multiple scales across different organ systems leading to an accumulation of damage that cannot be recovered through endogenous repair and regeneration. Biophysical, metabolic and immunological factors lead to a pro-fibrotic state, cellular senescence, and end-organ damage affecting both the heart and circulatory system.^3,4^ Ageing is driven by intrinsic processes that act at genetic, molecular and cellular targets, as well as extrinsic drivers such as lifestyle and environmental risk factors that modify these processes. In common with ageing in other organ systems, these mechanisms converge upon dysregulated inflammation, alteration of epigenetic modifications, and metabolic imbalances.^5^ A final common pathway of cardiovascular ageing is loss of tissue compliance which is manifest through diastolic dysfunction, interstitial fibrosis and vascular remodelling.^6,7^ Such changes can be assessed though non-invasive imaging and enable an estimate of how an individual’s cardiovascular system has aged relative to a normative population.^8^ An equivalent calculation of “age gap” has been shown to be a promising neuro-imaging marker for modelling dynamic lifespan trajectories of brain ageing.^9–11^

Here we used computer vision techniques to analyse cardiac magnetic resonance (CMR) imaging in 39,559 participants of UK Biobank to extract image-derived phenotypes of the three-dimensional (3D) geometry and motion of the heart as well as dynamic assessment of vascular function. These image-derived traits were used to train supervised machine learning algorithms to predict participants’ ages and derive a cardiovascular age-delta for each individual, quantifying the deviation in years from healthy ageing. Using age-delta as a trait for genome-wide common and exome-wide rare variant association analyses, we found ageing-associated loci containing genes known to be associated with tissue elasticity, myocardial contractility, inflammatory regulation and immune response to apoptosis. We also describe the relationship between cardiovascular risk factors and characteristics of premature cardiovascular ageing, and describe associated patterns of structural, functional and tissue-level changes in the heart. Together these analyses reveal the environmental and genetic mechanisms which underlie ageing of the cardiovascular system and indicate potential targets for risk modification.

## Results

### Study Overview

We analysed CMR data from 39,559 participants in UK Biobank using machine learning segmentation and motion tracking to measure multiple traits associated with cardiovascular structure, function, and fibrosis (Figure 1). Baseline characteristics of the population are shown in Extended Data Figure 1. A flow chart of analysis steps is depicted in Extended Data Figure 2. We first trained a machine learning model to define healthy cardiovascular ageing in a development set determined to be free of cardiac, respiratory and metabolic disease, using a gradient boosting algorithm (CatBoost).^12^ We used the CMR-derived traits to predict cardiovascular age and computed an age-delta for the difference between predicted age and chronological age. We then predicted cardiovascular age in the rest of the UK Biobank population (Figure 1c), and analysed the associations of the cardiovascular age-delta with traditional cardiovascular risk factors. We next performed a genome-wide association study (GWAS) of cardiovascular age-delta, having partitioned the data into discovery and validation sets by the release of data tranches by UK Biobank, and then a phenome-wide association study (PheWAS) to identify phenotypes associated with both age-delta and polygenic risk score (PRS) for age-delta. We performed rare variant association analyses using whole exome sequencing (WES). We also assessed any shared genetic architecture with age-deltas predicted using electrocardiographic (ECG) traits.

**Figure 1.**
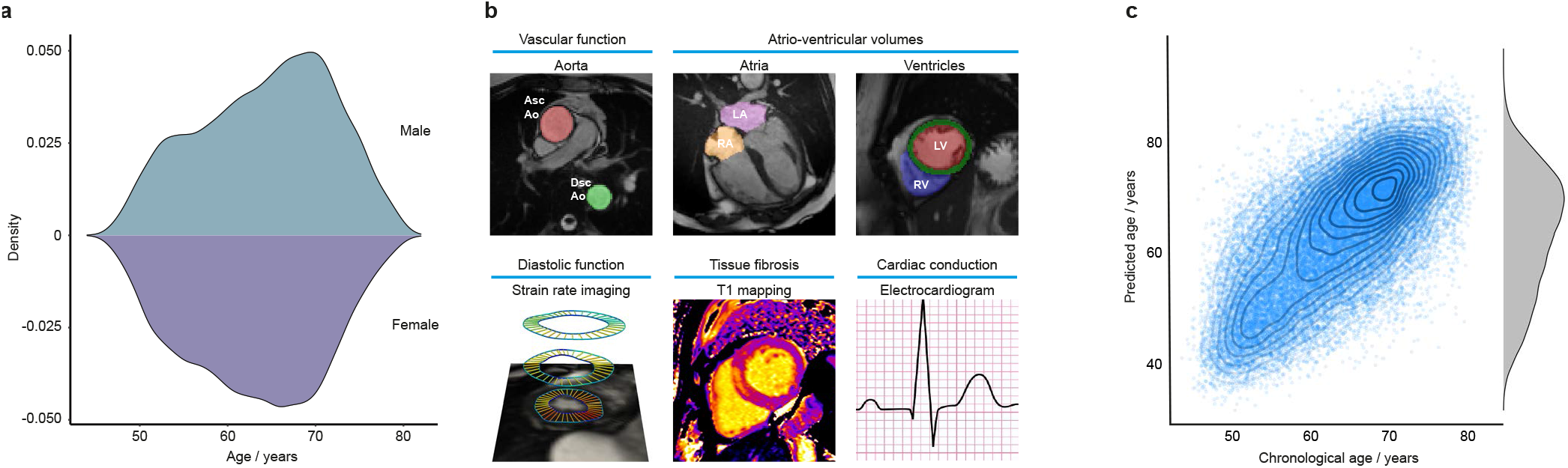
Summary of data used for cardiovascular age prediction in UK Biobank. **a**, Age distributions of participants by sex (kernel density estimates, 20,502 females, and 18,947 males). **b**, Imaging phenotypes used for prediction. The top row shows cardiac magnetic resonance images with automated time-resolved segmentation of the aorta and cardiac chambers. The bottom row shows an example of left ventricular motion analysis to derive radial strain rate and a parametric T1 map of the left ventricular myocardium. Resting electrocardiograms (ECGs) were used in an independent model for age prediction. Abbreviations: LA, left atrium; LV, left ventricle; RA, right atrium; RV, right ventricle; Asc Ao, ascending aorta; Dsc Ao, descending aorta. **c**, The relationship between predicted and chronological cardiovascular age (n=34,137, ages jittered, density contours, and a marginal density plot.)

### Image phenotyping

We performed automated quality-controlled analysis on CMR cine imaging to assess bi-atrial and bi-ventricular volumes and function, as well as left ventricular mass.^13^ Diastolic function, which is a key feature of the ageing heart, was assessed using motion analysis to derive end diastolic strain rates.^6^ We assessed diffuse myocardial fibrosis, an early feature of natural ageing,^14^ using native T1 mapping of the interventricular septum.^15^ Central vascular function was assessed by measuring aortic distensibility from central blood pressure estimates and dynamic aortic imaging.^13^ In total, 126 quantitative imaging phenotypes characterising structure, function and tissue characteristics were generated for each participant. To visualise variation in cardiac morphology with age we used time-resolved 3D morphometry of the heart.^16^

### Cardiovascular age prediction

A CatBoost machine learning model trained on healthy participants (n=4022), applied on a holdout test set (n=1044), yielded a coefficient of determination (R^2^) of 0.49, a Pearson correlation coefficient (|*r*|) between predicted age and chronological age of 0.70 (*P ≈* 0) and a mean absolute error (MAE) of 4.21 years. After bias correction, there was no correlation between cardiovascular age-delta and chronological age (|*r*| *<* 10^*−*16^, *P ≈* 1) showing that any deviations from healthy cardiovascular ageing were not related to the participants’ actual age.

The distribution and correlation between image-derived traits is shown in Extended Data Figures 3 and 4, and the feature importance of traits for cardiovascular age-delta is shown in Figure 2. Differences between sexes were observed across most phenotypes and were strongest for volumetric data. Sex was therefore used as a covariate in all analyses. There were also correlations between left and right chamber measurements, reflecting ventricular interdependence, with left-sided traits having greater predictive value. Arterial distensibility, in both the ascending and descending thoracic aorta, was the strongest predictor of age-delta. Late diastolic strain rates were also important contributors to the predictive model especially the radial component. 3D models of the heart also enabled visualisation of the morphological and functional changes related to age (Extended Data Figure 5). Left ventricular volumes decreased with age with remodelling of the lateral wall, while wall thickness showed progressive septal hypertrophy. Motion analysis demonstrated that regional changes in both left ventricular systolic contraction and diastolic relaxation occur with ageing.

**Figure 2.**
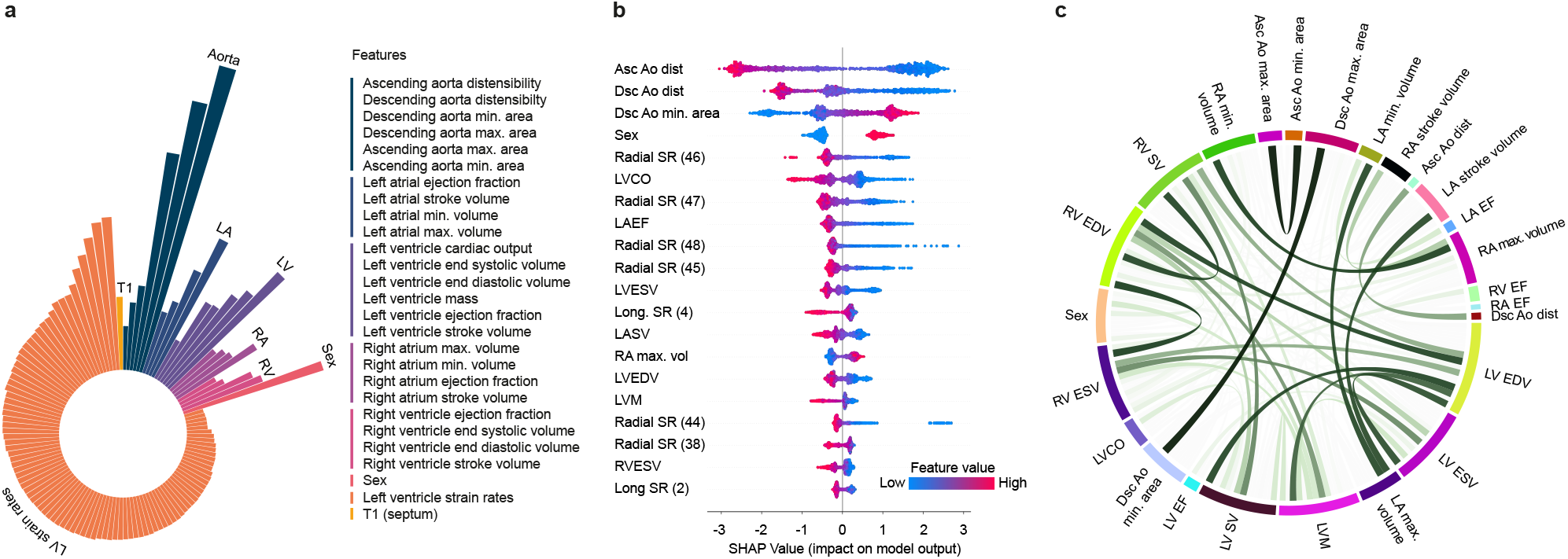
Features associated with cardiovascular ageing. **a**, Radial plot of log-transformed, normalised feature importance in cardiovascular age prediction using CatBoost grouped by category (aortic structure and function (Aorta), left atrial (LA) and left ventricular (LV) structure and function, right atrial (RA) and right ventricular (RV) structure and function, sex, left venticular strain rates and myocardial native septal T1. **b**, SHAP (Shapley Additive Explanations) plot of the top twenty features contributing to cardiovascular age prediction. The colour represents the feature value (red high, blue low), and its contribution to model prediction output. Features include ascending (Asc Ao) and descending aortic (Dsc Ao) distensibility (dist), descending aortic minimum cross-sectional area (Dsc Ao min. area), sex, radial and longitudinal strain rates (Radial SR, Long SR, numbers in bracket referring to frame number in cardiac cycle), left atrial stroke volume (LASV), left atrial ejection fraction (LAEF), left ventricular end systolic and diastolic volume (LVESV, LVEDV), left ventricular cardiac output (LVCO), left ventricular mass (LVM), right atrial maximum volume (RA max. vol) and right ventricular end systolic volume (RVESV). **c**, Circos plot of the correlation between imaging features. Ribbon widths are proportional to the absolute value of the Pearson correlation coefficient (|*r*|). For simplicity and clarity, absolute correlations are hidden where |*r*| < 0.4, and between radial and longitudinal strain measures. EDV, end diastolic volume; ESV, end systolic volume; EF, ejection fraction; SV, stroke volume. All plots n=34,137.

### Risk factors for cardiovascular ageing

A PheWAS of cardiovascular age-delta using PheCodes to aggregate a broad range of disease classifications in UK Biobank showed that this trait was predominantly associated with circulatory and metabolic disorders (Figure 3a). Using regression modelling of selected traits we showed that hypertension (+1.58 years, *P* = 2 *×* 10^*−*16^) and diabetes (+0.74 years, *P* = 0.000617) were both associated with an increase in predicted age (Figure 3b). Age-delta was also elevated in males with obesity (+0.46 years, *P* = 0.010639805), and females with coronary artery disease (+0.85 years, *P* = 0.035599339) and hypercholesterolaemia (+0.39 years, *P* = 0.048559275).

**Figure 3.**
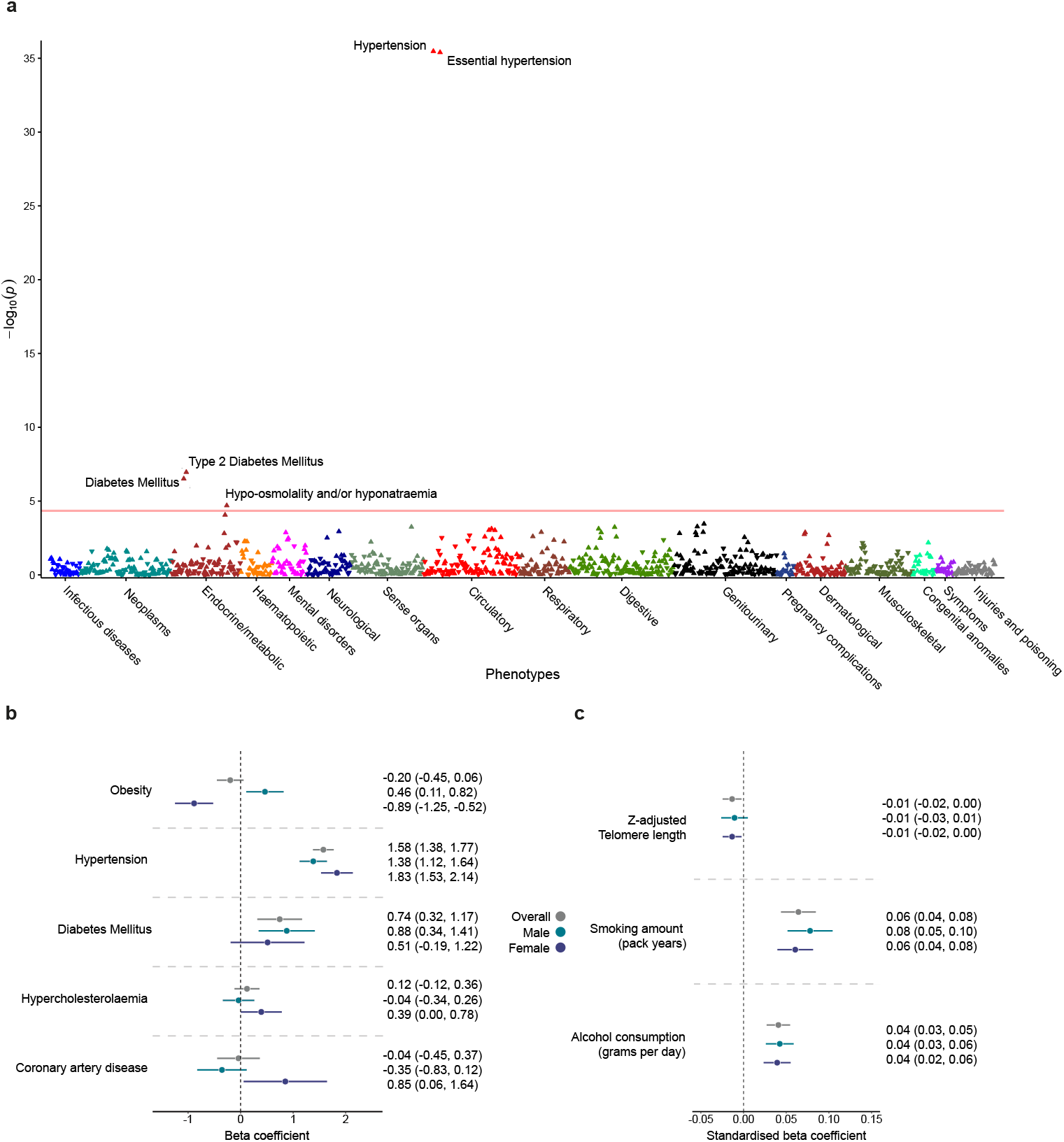
Associations of risk factors with cardiovascular age-delta. **a**, Phenome wide analysis of cardiovascular age-delta adjusted for age, age^2^, sex and the first ten genetic principal components by logistic regression (n = 34,137 participants). The red line represents the significance threshold after accounting for multiple testing (1149 phenotypes, *P <* 4.4×10*−*5). Upright triangles indicate positive correlations, and the inverted triangles indicate negative correlations. **b**, Linear regression analysis of categorical risk factors with cardiovascular age-delta. Data are presented as beta-coefficients point estimates, with 95% confidence intervals (CI), adjusted for age, age^2^ and sex, and compared with age and sex-matched controls. The test samples comprised n = 7089 participants with obesity, n = 7089 controls without obesity; n = 11,047 participants with hypertension, n = 11,047 controls without hypertension; n = 2466 participants with diabetes mellitus (DM), n = 2466 controls without DM; n = 7482 participants with hypercholesterolaemia, n = 7482 controls without hypercholesterolaemia; n = 2658 participants with coronary artery disease (CAD), n = 2658 controls without CAD. **c**, Linear regression analysis of quantitative risk factors with cardiovascular age-delta. Data are presented as standardised beta-coefficient point estimates (95% CI) per standard deviation (SD) increase in unit of risk factor (i.e. per SD pack year smoking, SD gram per day alcohol consumption and SD z-score telomere length increase), adjusted for age, age^2^ and sex. Data comprises 31,871 participants with telomere length data, 9283 participants with smoking data and 21,374 participants with alcohol consumption data.

Smoking (+0.031 years per smoking pack year, *P* = 7.0 *×* 10^*−*10^) and alcohol (+0.015 years per gram per day increase in daily alcohol consumption, *P* = 6.7 *×* 10^*−*9^) were also associated with adverse effects on cardiovascular ageing, with comparable effect sizes to brain ageing.^17^ Telomere length was associated with favourable effects on cardiovascular age (−0.10364 years per unit increase in z-adjusted telomere length, *P* = 0.017976312) (Figure 3c).

We found a modest association between cardiovascular age-delta and major adverse cardiovascular events (MACE) (hazard ratio (HR) 1.0861 (95% confidence intervals 1.0149 *−* 1.2124), *P* = 0.0222) when comparing upper and lower quartiles of age-delta in a covariate-adjusted model, but no significant associations in models considering cardiac age-delta as a continuous variable (see Supplementary Material for details).

To assess the potential use of age-delta as a surrogate marker of cardiovascular disease progression, we also examined the relationship between self-reported medication and cardiovascular age-delta. This showed that for most anti-hypertensives the effect on cardiovascular age was explained by measurable haemodynamic factors, while for beta-blockers, calcium channel blockers (CCBs) and metformin the association with cardiovascular age was independent of clinical risk factors, disease classes and haemodynamic parameters (see Supplementary Material).

### Genetic analyses of cardiovascular age-delta

#### Genome wide association study

All genetic analyses are reported in compliance with STREGA guidelines.^18^ The proportion of phenotypic variance in cardiovascular age-delta due to additive genetic variation (*h*^2^) explained by all genotyped single nucleotide polymorphisms (SNPs) was 10.5% (see Supplementary Material). We identified 5 genome-wide significant independent loci from our GWAS that associated with cardiovascular age-delta (*P* = 5 *×*10^*−*8^) (Figure 4). Summary information for the 5 loci identified using the full GWAS dataset are presented in Table 1, with further information provided in Supplementary Material. The nearest gene to the locus is defined, along with the most “likely gene” based on: evidence of a functional effect on a gene; previously documented cardiovascular disease association; or reported mechanism potentially involved in cellular ageing processes (Supplementary Material). The lead variants have known roles in myocardial contractility (Titin, *TTN*),^19^ and arterial mechanics (Elastin, *ELN*),^20^ and have been implicated in pro-inflammatory activity and antihypertensive responses (Phospholipase C Epsilon 1, *PLCE1*).^21,22^ Additionally, *TTN* was a likely causal gene for ECG age-delta amongst 7 other independently-associated loci that include genes related to trabecular development (T-Box Transcription Factor 3, *TBX3*),^23^ and regulation of cardiac rhythm (Sodium Voltage-Gated Channel Alpha Subunit 5, *SCN5A*; Calcium/Calmodulin Dependent Protein Kinase II Delta, *CAMK2D* and Myozenin 1, *MYOZ1*) (see Supplementary Material).

**Table 1.**
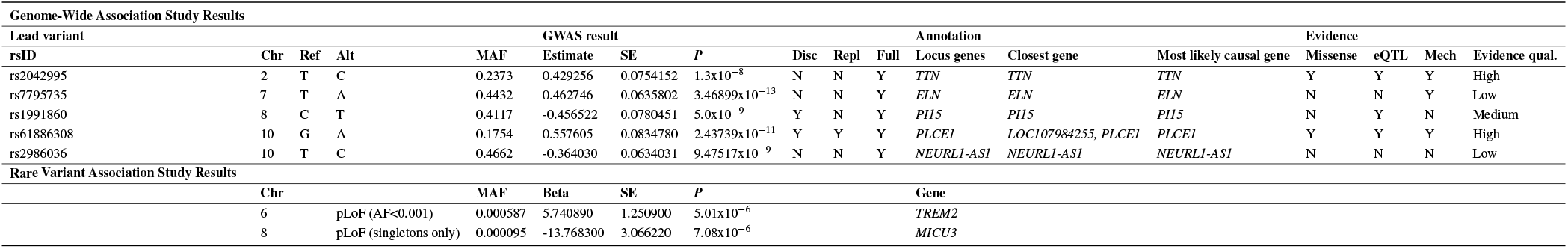
Summary of genetic association studies.. Summary information on the lead single nucleotide polymorphisms (SNPs) of the genome wide association study (GWAS)-identified significant loci and the genes identified from the rare variant association study (RVAS). For each significant locus, lead SNP summary information is provided (Chr, chromosome; Ref, reference allele; Alt, alternative allele; MAF, minor allele frequency), with GWAS summary statistics (Estimate, beta coefficient; SE, standard error; *P, P*-value). Details of which set of the data that the lead SNP reached genome-wide significance in is also presented (Disc, discovery set; Repl, replication set; Full, full dataset). Variant to gene annotation is provided and a summary of strength of evidence of gene mapping. Abbreviations: Missense (missense variant); eQTL status (co-localisation of GWAS signal with an expression quantitative trait loci for the gene in a plausible tissue type); Mech (plausible mechanistic link between the gene and the phenotype i.e. ageing); and Evidence qual. (Quality of evidence of variant to gene mapping, strength of evidence deemed “high”, “medium” or “low”.). For RVAS, the MAF indicates the frequency of observing pLoF variants in the specified mask. Analysis performed on the following datasets: discovery, n=20,058; validation, n=9448; and full, n=29,506. Effect estimate, standard error and *P*-values relate to results of the full GWAS.

**Figure 4.**
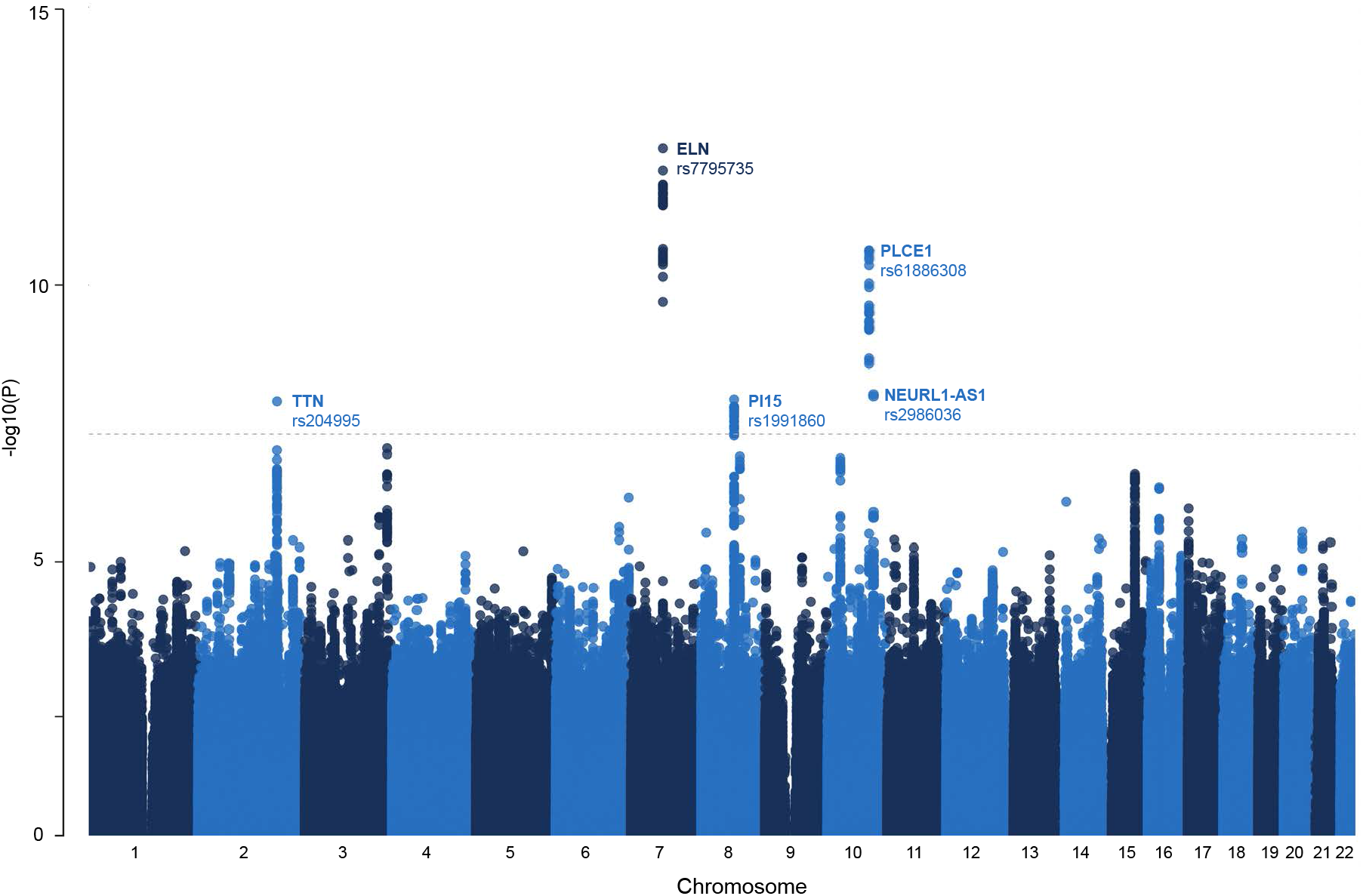
Manhattan plot with the results of the genome wide association study of cardiovascular age-delta. This figure shows the −*log*_10_(*P* value) on the *y* axis across all autosomal chromosomal positions (*x* axis). The dotted line indicates genome-wide significance (*P* = 5 *×*10*−*8, n = 29,506). Significant loci are labeled by their most likely causal gene and the lead single nucleotide polymorphism (Table 1).

A covariate-corrected polygenic risk score (PRS) comprising 13 SNPs selected using a clumping/thresholding approach for cardiac age-delta was evaluated in 373,948 independent genotyped participants of UK Biobank, and showed an association with hypertension (*P* = 2.9 *×*10^*−*12^) (see Supplementary Material).

#### Rare variant association study

Predicted loss of function variant (pLoF) gene burden testing using Regenie,^24^ with three variant prioritisation masks on allele frequencies applied, identified two further genes associated with cardiac ageing independent of common variant signals. These were Triggering Receptor Expressed On Myeloid Cells 2 (*TREM2*) and Mitochondrial Calcium Uptake Family Member 3 (*MICU3*). *TREM2* encodes a transmembrane receptor expressed in the central nervous system and macrophages, and has been linked to anti-inflammatory effects in atherosclerotic plaque, cardiac tissue repair and immunomodulation of monocytes in the heart.^25–27^ *MICU3* encodes a uniporter channel in the heart and skeletal muscle, which regulates calcium homeostasis.^28,29^

## Discussion

Cardiovascular ageing is an interaction of genetic, cellular, and biophysical processes that results in declining adaptive homeostasis. In this study, we used machine learning of multiple image-derived cardiovascular phenotypes to predict biological age and determine the genetic and environmental associations with deviation from healthy ageing.

We found that healthy ageing was associated with a progressive decline in both vascular and myocardial tissue compliance, and these traits were also the strongest predictors of deviation from healthy cardiovascular ageing. A decline in diastolic function is recognised as a hallmark of cardiac ageing, which occurs through multiple profibrotic and energetic pathways.^30,31^ A key driver of diastolic impairment is myocardial interstitial fibrosis and we found that an imaging biomarker of fibrosis predicted accelerated ageing. Variation in this specific biomarker is thought to be mediated by glucose metabolism, tissue repair and oxidative stress,^32^ with fibrosis being a final common pathway after homeostatic mechanisms are exhausted.^33^ Three dimensional image analysis showed that ageing results in asymmetrical remodelling, and patterns of contractility and relaxation in the left ventricle. We observed progressive septal hypertrophy which is recognised as an independent feature of ageing in longitudinal studies.^34^ The aorta also becomes stiffer with age, which is associated with numerous cardiovascular disease endpoints.^35^ Elastin fragmentation, endothelial dysfunction and deposition of advanced glycation end-products are thought to play a causal role,^36^ and distensibility is associated with pathways related to cardiovascular development, extracellular matrix production, and smooth muscle cell contraction.^37^

Our study provides insights into the biological basis of heterogeneity in cardiovascular ageing. One of our loci implicated *PLCE1* which regulates contractile myocardial reserve with loss of expressed PLC*ε* signaling sensitizing the heart to the development of hypertrophy in response to chronic cardiac stress.^38^ PLC*ε* also regulates inflammatory responses to myocardial injury,^21^ consistent with the putative causal relationship of “inflammageing” with cardiovascular disease and multimorbidity.^39^ *ELN* was also a leading locus with elastin being an abundant extracellular matrix protein that provides elasticity and resilience to tissues including the distensibility of blood vessels. The low turnover of elastin and chronic biophysical stress makes it susceptible to age-related changes caused by mechanical fracture, proteolysis, and calcification.^20^ Age-associated arterial structural and functional decline may not be inevitable and lifestyle interventions have been shown to preserve vascular elasticity.^40^ We identified common variants in a cardiomyopathy-associated gene (*TTN*) suggesting a role for sarcomere homeostasis as a mediator of human cardiac ageing. This may occur through an age-dependent effect on adaptive capacity that is distinct to the changes in titin isoform expression seen in heart failure.^41^ Common variants in *TTN* were also implicated in ECG age-delta suggesting a shared genetic mechanism with conduction-associated traits. Using WES we found rare variants in two further genes that were were associated with deviation from healthy cardiovascular ageing. The protein expressed by *TREM2* is an immune regulator controlling monocyte/macrophage transitions in response to apoptosis in the heart.^26^ This suggests it may play a role at the interface of immunomodulation and cardioprotection in the ageing heart. *MICU3* plays an important role in regulating pathological calcium overload in the heart,^28^ and has been implicated in determining antioxidant responses in skeletal muscle ageing.^29^

Overall the SNP-based heritability of cardiovascular ageing was 10.5% suggesting that non-genetic contributions predominantly affect ageing. We found that cardiometabolic risk factors including hypertension, diabetes and hypercholesterolaemia all contribute to an increased age-delta. “Early vascular ageing” is thought to be induced by the integrated effect of haemodynamic factors, glycaemic dysregulation and fetal programming,^42^ while diabetes and dyslipidaemia have also been shown to be causally associated with diastolic dysfunction which is also a major contributor to premature ageing in the heart.^6^ Obesity showed divergent effects in each sex and in men was associated with accelerated ageing while the opposite effect was observed in women. Obesity and overweight are associated with a non-linear increased risk of all cause mortality,^43^ but misclassification of body composition, particularly among women, may lead to biased risk estimates.^44^ There is evidence that obesity accelerates the ageing process through epigenetic alterations, mitochondrial dysfunction, cellular senescence and a pro-inflammatory state.^45^ Obesity may also affect telomere dynamics and accelerate the ageing process although the data is heterogeneous.^46^ We did observe an association between telomere shortening and premature cardiovascular ageing although the effect size was relatively small. Alcohol and smoking showed adverse associations with cardiovascular ageing that were of similar effects sizes to those observed in brain ageing.^17^ For cardiovascular age-delta to be a useful surrogate endpoint it would need to be correlated with hard clinical outcomes in the absence of intervention and modulated by established therapies, with its magnitude of modulation related to the effect of the intervention.^47^ In cross-sectional analyses, whilst most antihypertensive effects on ageing were explained by known risk factors, beta-blockers, CCB, and metformin were independent predictors of age-delta. Further research is needed to investigate response of cardiac age-delta to drug therapy, either in appropriately designed interventional studies, or through analysis of emerging observational data, whilst appropriately accounting for potential biases in this type of data.

There are limitations of this study. The rate of participation in the UK Biobank is higher among women, older age groups, and persons living in less socioeconomically deprived areas.^48^ The population is predominantly European and further work is required to explore ageing traits and outcomes in people of diverse ancestries and social groups as an accelerated ageing phenotype may be observed due to the interaction of biological, psychosocial, socioeconomic factors.^49^ Cardiovascular age-delta is derived at a single time-point and we could not assess within-person ageing of the cardiovascular system. Longitudinal imaging would also help to determine the relative contribution of early-life influences and polygenic risk compared to accelerated ageing in response to external risk factors in adult life.

In conclusion, we found that cardiovascular ageing is linked to multiple modifiable risk factors, shows distinct patterns of remodelling, and is associated with loci related to genes regulating sarcomere homeostasis, myocardial immunomodulation, and tissue responses to biophysical stress.

## Methods

All analyses in this study are available online (https://github.com/ImperialCollegeLondon/cardiovascular_ageing) and were conducted with R v.>3.6.0,^50^ and Python v.3.9.^51^

### Participants

UK Biobank comprises approximately 500,000 community-dwelling participants aged 40–69 years who were recruited across the United Kingdom between 2006 and 2010.^2^ All participants provided written informed consent for participation in the study, which was approved by the National Research Ethics Service (11/NW/0382). Our study was conducted under terms of access approval numbers 28807 and 40616. A range of available data were included in this study comprising genotyping arrays and WES, cardiac imaging, health-related diagnoses, and biological samples.

There were 488,252 genotyped participants of which 454,787 have WES. We partitioned 39,559 participants with both CMR and genotyping array data into two tranches by date of release from UK Biobank providing a discovery dataset of 26,893 participants and a validation dataset of 12,666 participants. We generated age predictions in a subset of 20,058 discovery and 9448 European ancestry participants.

### Non-imaging phenotypes

Participants underwent a resting 12 lead electrocardiogram. Other phenotypes were collected by touch screen questionnaire, interview, biophysical measurement, hospital episode statistics, and primary care data (see Supplementary Material). Leucocyte telomere length (LTL) was measured using a multiplex qPCR assay from 488,415 available DNA samples of participants in UK Biobank.^52^ After quality control (QC), valid LTL measurements were available for 472,577 individuals and log-transformed and z-standardised values were used for analysis (UK Biobank data field code 22192). Details of how each phenotype was acquired are available on the UK Biobank Showcase (http://biobank.ctsu.ox.ac.uk/crystal/).

### Imaging protocol

A standardised CMR protocol was followed to acquire two-dimensional, retrospectively-gated cine imaging on a 1.5T magnet (Siemens Healthineers, Erlangen, Germany).^53^ Short-axis plane cine imaging involved acquiring a contiguous stack of images from left ventricular base to apex, and long axis cine imaging was also performed in the two and four chamber views. Cine sequences all consisted of 50 cardiac phases with an acquired temporal resolution of 31 ms.^53^ Transverse cine imaging of the ascending and descending thoracic aorta was also performed. Native T1 mapping within a single breath hold was performed at mid-ventricular level using a shortened modified Look-Locker inversion recovery (ShMOLLI) sequence. Imaging phenotypes all underwent QC prior to use in analysis.^54^

### Cardiac image analysis

As previously described,^6^ automated segmentation of the short-axis and long-axis cine images in UK Biobank was performed using fully convolutional networks. Image segmentation in the UK Biobank dataset using this deep learning network is equivalent to expert human readers.^54^ Volumes (end-diastolic, end-systolic, and stroke volume) and ejection fraction were determined for both ventricles. Myocardial volumes were used to compute left ventricular myocardial mass assuming a density of 1.05 g.ml ^−1^. Atrial volumes were calculated using the biplane area–length formula 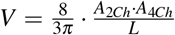(where *A*_2*Ch*_ and *A*_4*Ch*_ are the atrial areas on the two and four-chamber cine views respectively, and *L* is the averaged longitudinal diameter across two views). Measurements were indexed to body surface area (BSA) as per the Du Bois formula: 0.20247 *∗* (*Weight*^0.425^) *∗* (*Height*^0.725^), with weight in kg and height in m. The heart was divided into 16 standardised anatomical segments, excluding the true apex.^55^

The aorta was segmented using the cine images with a spatio-temporal neural network,^56^ from which maximum and minimum cross-sectional areas were derived. Distensibility was calculated using central blood pressure estimates obtained using peripheral pulse-wave analysis (Vicorder, Wuerzburg, Germany).^13^

Non-rigid image registration between successive frames enabled motion tracking on greyscale images.^57^ Registration errors were minimised by tracking motion in both backwards and forwards directions from end-diastole, resulting in an averaged displacement field,^13^ which was then used to warp segmentations from end-diastole to successive adjacent frames.*L*_*dir*_Circumferential (*E*_cc_) and radial (*E*_rr_) strains were calculated using short axis cines as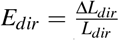, where *dir* represents circumferential or radial direction, *L*_*dir*_ the absolute length of a line segment along this direction and Δ*L*_*dir*_ its change in length over time. Longitudinal (*E*_ll_) strain was calculated from long-axis four-chamber motion tracking measured at basal, mid-ventricular, and apical levels. Segmental and global peak strains were then calculated. Strain rate was computed as the first derivative of strain and thereafter peak early diastolic strain rate in radial (PDSR_rr_) and longitudinal (PDSR_ll_) planes then identified. The ShMOLLI T1 maps were analysed using probabilistic hierarchical segmentation with automated quality control defining a region of interest within the interventricular septum as previously validated.^15^ Blood pool T1 was used as a linear correction of myocardial T1 values.^15,58^

Three dimensional visualisation of left ventricular shape and motion with respect to age was performed through atlas-based registration of the segmentations and motion fields. We calculated the mean phenotypes (shape, wall thickness and motion pathlines) for each decade of chronological age and represented these values on the epicardial surface of the models relative to participants aged 40-49 years.^16^

### Cardiovascular age prediction

To define a model of healthy cardiovascular ageing, as previously validated in other organ systems,^11,59–61^ we first partitioned the 39,559 participants into a development set, which consisted of 5065 “healthy” individuals that were free of cardiac, metabolic or respiratory disease, with a body mass index below 30 (see Supplementary Material for details). We randomly split this group into separate training (80%, n = 4021) and test (20%, n = 1044) sets.

All features were normalised through scaling and mean-centering of data. We used CatBoost, a decision-tree based gradient boosting machine learning algorithm, on this feature set to predict the age of each participant in the healthy training and test sets. CatBoost was trained using the default hyperparameter set, except for early stopping rounds (patience) between 50 and 100, which was chosen by the default hyperparameter search method. A 10% validation holdout set (n = 403) was used for hyperparameter search (using the Python package Optuna). From the remaining training dataset, an additional 10% (n = 362) holdout set was taken to be used by CatBoost for internal early stopping evaluation (therefore, training CatBoost with the remaining n = 3256 instances). Thirty models (with distinct random initialization seeds) were trained and the one with the lowest mean absolute error on the holdout dataset was chosen.

The trained CatBoost model was used to predict age in participants from the evaluation set (n=34,147).^10,11^ As in brain age modelling,^11,17,62–64^ we corrected for the correlation between age-delta and chronological age. For this bias-correction we used linear regression between the initial uncorrected cardiac age-delta against chronological age.^63^ An offset is then calculated by multiplying chronological age by the slope of the regression line, and adding the intercept. A corrected predicted age is computed by subtracting this offset from the uncorrected predicted age.

### Evaluating associations of cardiovascular age-delta with risk factors

To explore associations of cardiovascular age-delta with classical cardiovascular risk factors, we curated five groups from the evaluation set, comprising individuals with obesity, hypertension, diabetes mellitus (DM), hypercholesterolaemia and coronary artery disease. We additionally obtained smoking frequency and z-adjusted leukocyte telomere length data, and derived alcohol consumption for participants (see Supplementary Material for details). For each of the five diseases, we selected a set of controls from the test set who did not have the disease, matched for age and sex using 1:1 propensity score matching. Each test sample thus had the same number of controls as number with the disease of interest, and data from some participants also acted as control data for several disease groups (see Supplementary Methods for further detail). Consistent with recommendations from brain ageing literature, all onward statistical analyses adjusted for age, age^2^ and sex.^10,64^ We then fit a linear regression model to test for the association between cardiovascular age-delta and presence of disease. For continuous risk variables (alcohol and smoking consumption, and telomere length), we fit a linear regression model in all individuals of the test data set that had complete data for these variables to assess the association between cardiovascular age-delta and risk factor. For modelling associations with self-reported medication see Supplementary Materials.

### Outcome analysis

For assessing the association of cardiovascular age-delta with prospective cardiovascular events, we calculate time-to-first major cardiovascular event (MACE) (stroke, heart failure, arrhythmia, cardiovascular death; see Supplementary Material for details) from the time of the imaging visit. We stratify by MACE event prior to the MRI visit. Participants are split into quartiles and associations with MACE events are assessed through both descriptive analysis (cumulative incidence curves with all-cause death as a competing event) and model-based analysis with both a fully covariate-adjusted Cox model (age, sex, obesity, diabetes, smoking, alcohol consumption, hypertension, coronary artery disease, hypercholesterolaemia) or a minimally covariate-adjusted Cox model (age, sex).

### Genotyping and sample quality control

Genotyping of UK Biobank participants has been detailed previously,^65^ and in brief, UK Biobank genotyping for 488,252 participants was performed on UK BiLEVE or UK Biobank Axiom arrays and imputation performed with the HaplotypeReference Consortium panel and the UK10K+1000 Genomes Project panel. We used UK Biobank Imputation V3 (in GRCh37 coordinates). The UK Biobank released whole exome data sequencing of 454,787 participants in 2021, and details regarding sequencing methods and variant calling procedures are described elsewhere.^66^ We utilised genotypes in their released PLINK-format files, and restricted the cohort to the European population. We performed standard quality control steps recommended by UK Biobank, detailed in the Supplementary Material.

### GWAS analysis

GWAS analyses for cardiovascular age-delta was performed with PLINK (v.2) in the discovery (n=20,058), validation (n=9448) and full (n=29,506) cohorts. All GWAS analyses were adjusted for sex, age (at time of MRI), age^2^, the first ten genetic principal components, MRI assessment centre and genotyping array. Post-GWAS analysis removed SNPs with a Hardy–Weinberg equilibrium *P <*0.05 and minor allele frequency (MAF)*<*0.005. Lead variants for each locus were assigned likely causal genes using variant annotations. Expression quantitative trait loci (eQTL) evidence for each locus was extensively searched for using the GTEx Portal and where available, full summary statistics were downloaded to assess co-localisation (Supplementary Material).

### Identification of rare variant gene-based associations that were independent of common variant signals

We performed rare-variants burden testing on UK Biobank WES using Regenie^24^ on the Research Analysis Platform (RAP) (https://ukbiobank.dnanexus.com). The intersection of European participants with exome sequencing data, the age-delta phenotype, and array data after QC is 31,515. We further quality-checked the WES data by requiring that at least 90% of genotypes for a given variant, independent of variant allele zygosity, have a read depth of at least 10 (the 90pct10dp filter) to avoid spurious hits. After QC, we ran step 1 to obtain predictors of individual trait values based on common genetic data, which were then used in step 2 for the rare variant gene burden testing.

For step 2, we annotated pLoF variants and defined gene variant sets using the UK Biobank 450K Exome helper files,^67^ also see UK Biobank Resource 916. We tested the pLoF variants on three separate burden masks per gene, based on the frequency of the alternative allele of the variants: MAF*≤*1%, MAF*≤*0.1% and singletons only. We tested genes with enough (5) pLoF carriers across all samples. Namely, the number of genes tested for singletons is 4271, for MAF*≤*0.1% is 10431, and for MAF*≤*1% is 10592. We performed Regenie step 2 with the adjustment of sex, age (at time of MRI), age^2^, the first ten genetic principal components, and genotyping array. The association was considered significant after multiple testing correction at *α* = 0.05.

### Polygenic risk score (PRS) and PheWAS

Candidate variants for PRS for the cardiovascular age-delta were obtained based on the respective GWAS results by performing clumping (PLINK v1.9) using a linkage disequilibrium (LD) threshold of *R*^2^ = 0.1 (in a window of 250kb) and considering all SNPs with *P <* 0.001. These selected variants were then used to construct a genetic score for all individuals in the dataset using linear scoring in PLINK v2. Missing genotypes were imputed using the default mean imputation approach. We have additionally constructed a multivariable linear model evaluated on the European subset of the full imaging cohort, using sex, age (at time of MRI), age^2^, the first ten genetic principal components, MRI assessment centre and genotyping array as additional covariates, and cardiovascular age-delta as the dependent variable. We report the variance explained by the PRS as the difference of linear regression *R*^2^ between a model of age-delta with all non-genetic covariates and the model that additionally includes the PRS as a covariate (see Supplementary Material).

PheWAS of imaging-derived cardiovascular age-delta and age-delta PRS was performed in European ancestry participants in the UK Biobank. PRS PheWAS was performed in participants that were not included in the derivation GWAS (*n* = 373,948), whilst imaging-derived age-delta PheWAS was performed in the same imaging cohort (*n* = 34,137). For phenotypes with at least 20 cases, association was tested using logistic regression, adjusting for age, age^2^, sex, and first ten genetic principal components. Statistical significant threshold was adjusted for the total number of phenotypes tested (1149 phenotypes), and data presented with Manhattan plots grouped by body systems. PheWAS was performed using the PheWAS package in R version 4.0.3.

### Age prediction using resting electrocardiograms

Cardiac age-delta predictions have been performed previously using electrocardiogram (ECG) data.^68^ We were interested to see how predictions from CMR imaging and resting ECG data might correlate, and also investigate shared genetic variants for age-delta. To achieve this, we adapted a previously published neural network model to perform the ECG-based predictions. We trained the model using ECG input data from the UK Biobank RAP and re-formatted these according to the requirements of the model. Subsequently, we fine-tuned the pre-trained model using the same development set used in the imaging based predictions. Details on data input adaption and model refinement are provided in Supplementary Material. We used the same statistical and GWAS approach for ECG age-delta as described for image phenotype-derived cardiovascular age-delta.

## Data Availability

The analysis code is freely available on GitHub (https://github.com/ImperialCollegeLondon/cardiovascular_ageing).
All raw and derived data in this study are available from UK Biobank (http://www.ukbiobank.ac.uk).

## Acknowledgments

The study was supported by Bayer AG; Medical Research Council (MC_UP_1605/13); National Institute for Health Research (NIHR) Imperial College Biomedical Research Centre; British Heart Foundation (RG/19/6/34387, RE/18/4/34215); and the Sir Jules Thorn Charitable Trust [21JTA]. This research has been conducted using the UK Biobank Resource under Application Numbers 28807 and 40616. The genetic association analyses were conducted on the UK Biobank Research Analysis Platform (https://ukbiobank.dnanexus.com). We thank James Cole (UCL) and Tobias Kaufmann (University of Tübingen) for advice on age-delta modelling. We also thank Antonio de Marvao, Tim Dawes, Ghalib Bello, and Carlo Biffi (Imperial College London) for developing the three dimensional cardiac modelling used in this study. We also acknowledge Esther Puyól-Anton, Bram Ruijsink and Reza Razavi for the T1 mapping data.

For the purpose of open access, the authors have applied a creative commons attribution (CC BY) licence to any author accepted manuscript version arising.

## Code Availability

The analysis code is freely available on GitHub (https://github.com/ImperialCollegeLondon/cardiovascular_ageing).

## Data Availability

All raw and derived data in this study are available from UK Biobank (http://www.ukbiobank.ac.uk/). GWAS summary level data are publicly available through the GWAS catalogue.

## Author contributions

M.S. and M.H.de A.I. performed the formal analyses and co-wrote the paper; W.B. and A.P.K. performed the image analysis; K.A.McG., E.E, I.K., C.L., J.S.W., J.M., C.B. and D.F.F. performed or interpreted the genetic and outcome analyses; S.L.Z., P-R.S. and A.C. performed additional data modelling; M.R.W critically revised the manuscript; D.P.O’R conceived the study, managed the project and revised the manuscript. All authors reviewed the final manuscript.

## Extended Data Figures

**Extended Data Figure 1.**
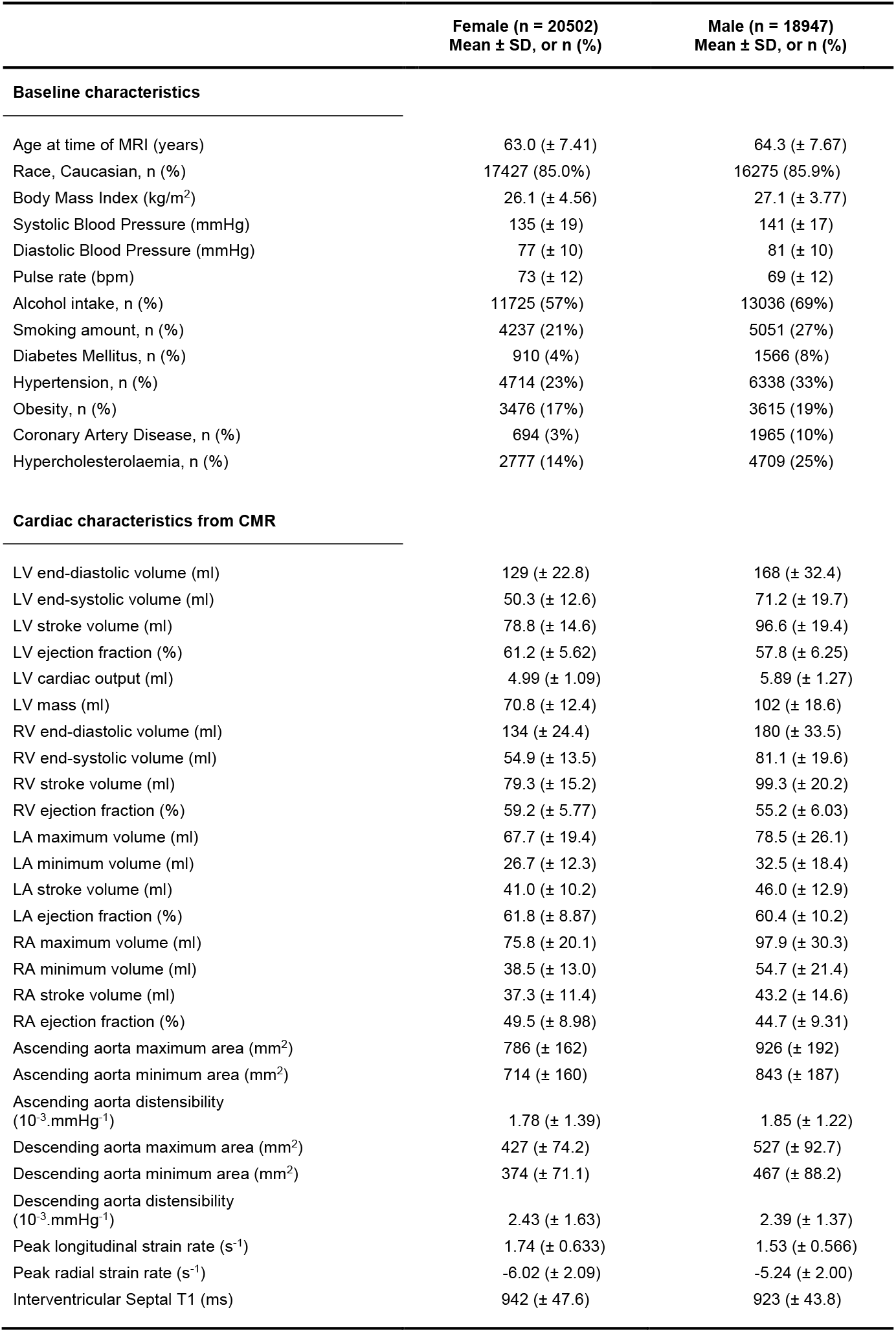
Baseline participant characteristics. A summary of clinical and imaging characteristics in the UK Biobank population stratified by sex. Abbreviations: LA, left atrium; LV, left ventricle; RA, right atrium; RV, right ventricle; T1, longitudinal relaxation time of tissue.

**Extended Data Figure 2.**
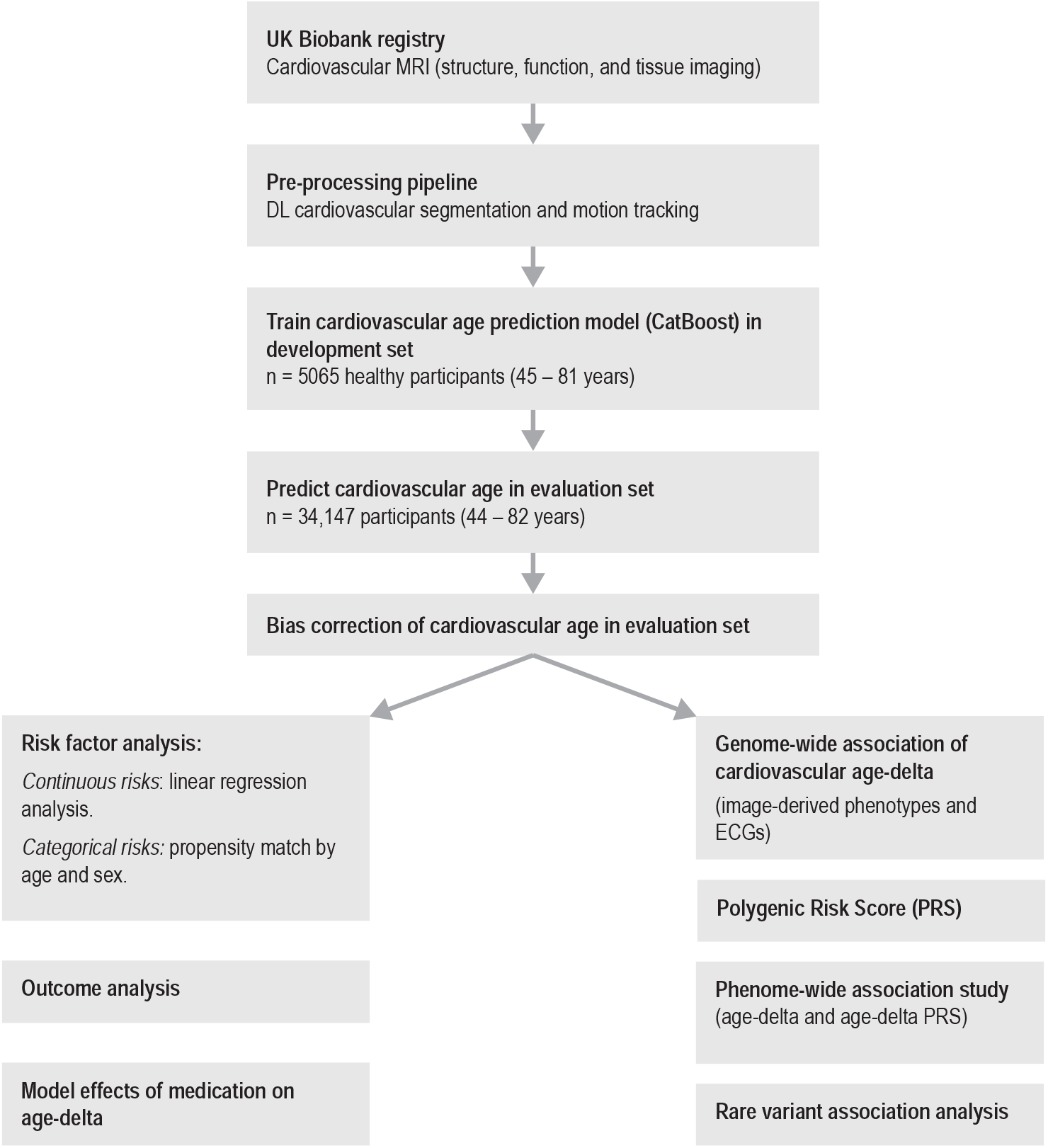
Study flowchart. A summary of the main steps in our analysis of cardiovascular imaging traits, prediction of age-delta and genetic/environmental associations. MRI, Magnetic Resonance Imaging; DL, Deep Learning; ECG, electrocardiogram

**Extended Data Figure 3.**
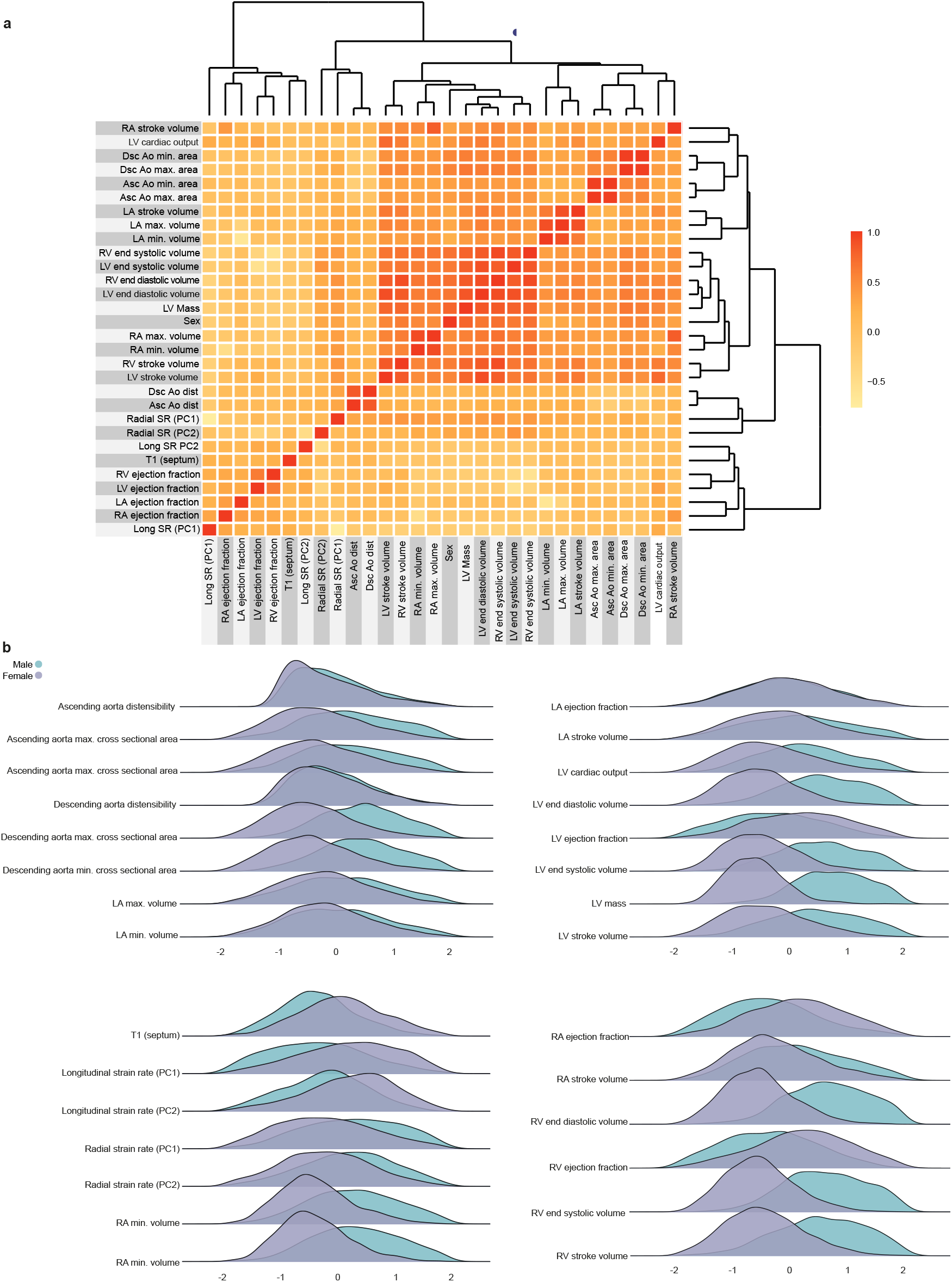
A summary of non-imaging and imaging features used in cardiovascular age prediction model. **a**, Heatmap for the features, with colours representing the Pearson correlation coefficient (n=39,559). **b**, Ridge plots summarising the distribution densities of features, normalised for visualisation purposes (n=39,559). Asc Ao dist, ascending aortic distensibility; Asc Ao min./max. area, ascending aortic minimum/maximum cross-sectional area; Dsc Ao dist, descending aortic distensibility; Dsc Ao min./max. area, descending aortic minimum/maximum cross-sectional area; LA, left atrium; LASV, left atrial stroke volume; LAEF, left atrial ejection fraction; LVESV, left ventricular end systolic volume; LVEDV, left ventricular end diastolic volume; LVCO, left ventricular cardiac output; LVM, left ventricular mass; LV, left ventricle; PC, principal component; RA, right atrium; RV, right ventricle; Radial/Long SR, radial/longitudinal strain rates (numbers in bracket referring to frame number in cardiac cycle); RA max. vol, right atrial maximum volume; RVESV, right ventricular end systolic volume.

**Extended Data Figure 4.**
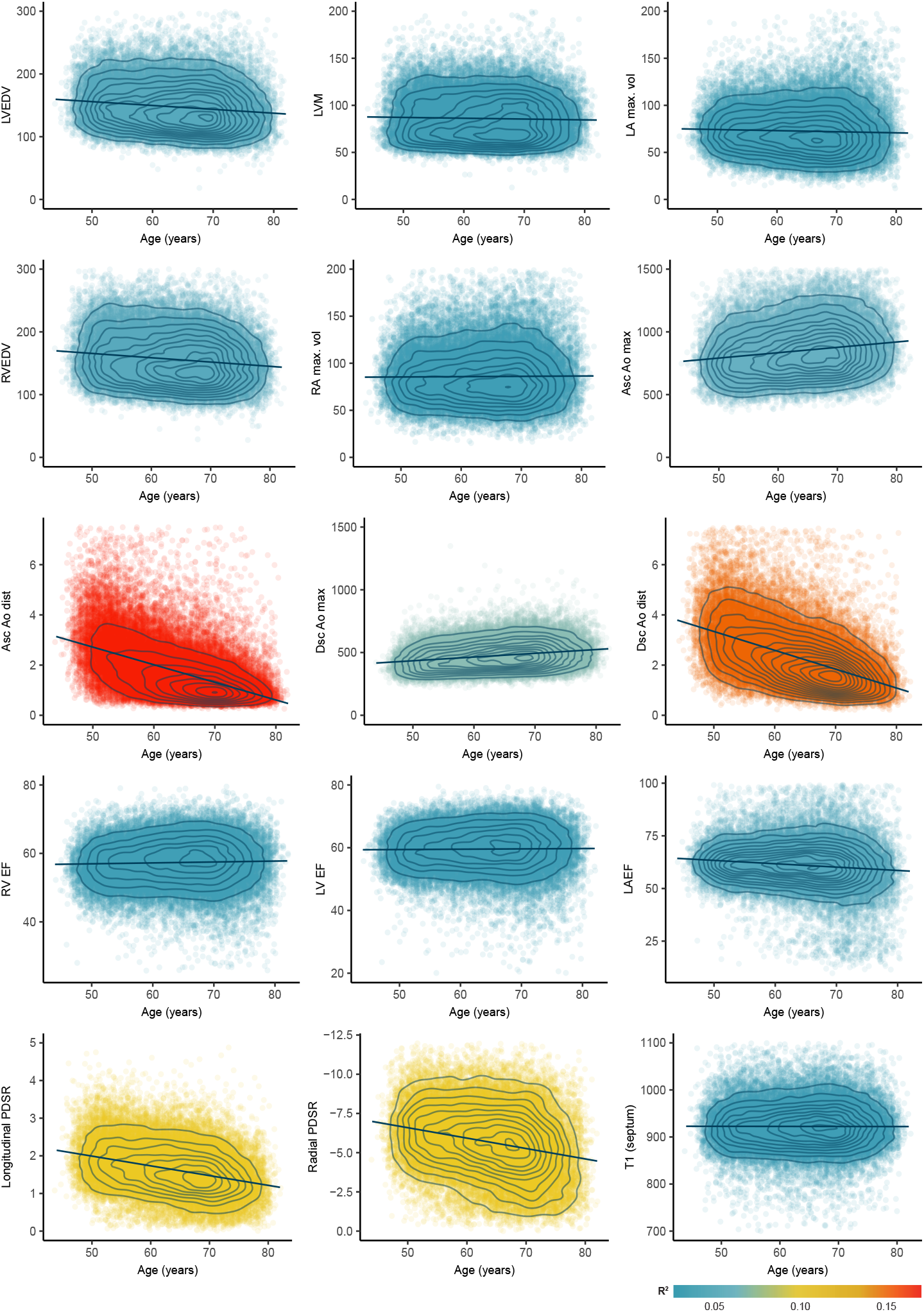
Image phenotype associations with chronological age. In total we measured 126 image-derived phenotypes including temporal motion analysis of cine imaging. A selection of 15 representative phenotypes of volumes, function, and tissue characterisation are shown with their relationship to chronological age at the time of imaging. (n=39443, ages jittered, density contours, point colours represent coefficient of determination (R^2^)). LVEDV, left ventricular end diastolic volume; LVM, left ventricular mass; LA max. vol, left atrial maximum volume; RVEDV, right ventricular end diastolic volume; RA max. vol, right atrial maximum volume; Asc/Dsc Ao max., ascending/descending aortic maximal cross-sectional area; Asc/Dsc Ao dist., ascending/descending aortic distensibility; RV/LVEF, right/left ventricular ejection fraction; LAEF, left atrial ejection fraction; PDSR, peak diastolic strain rate; T1,longitudinal relaxation time of tissue.

**Extended Data Figure 5.**
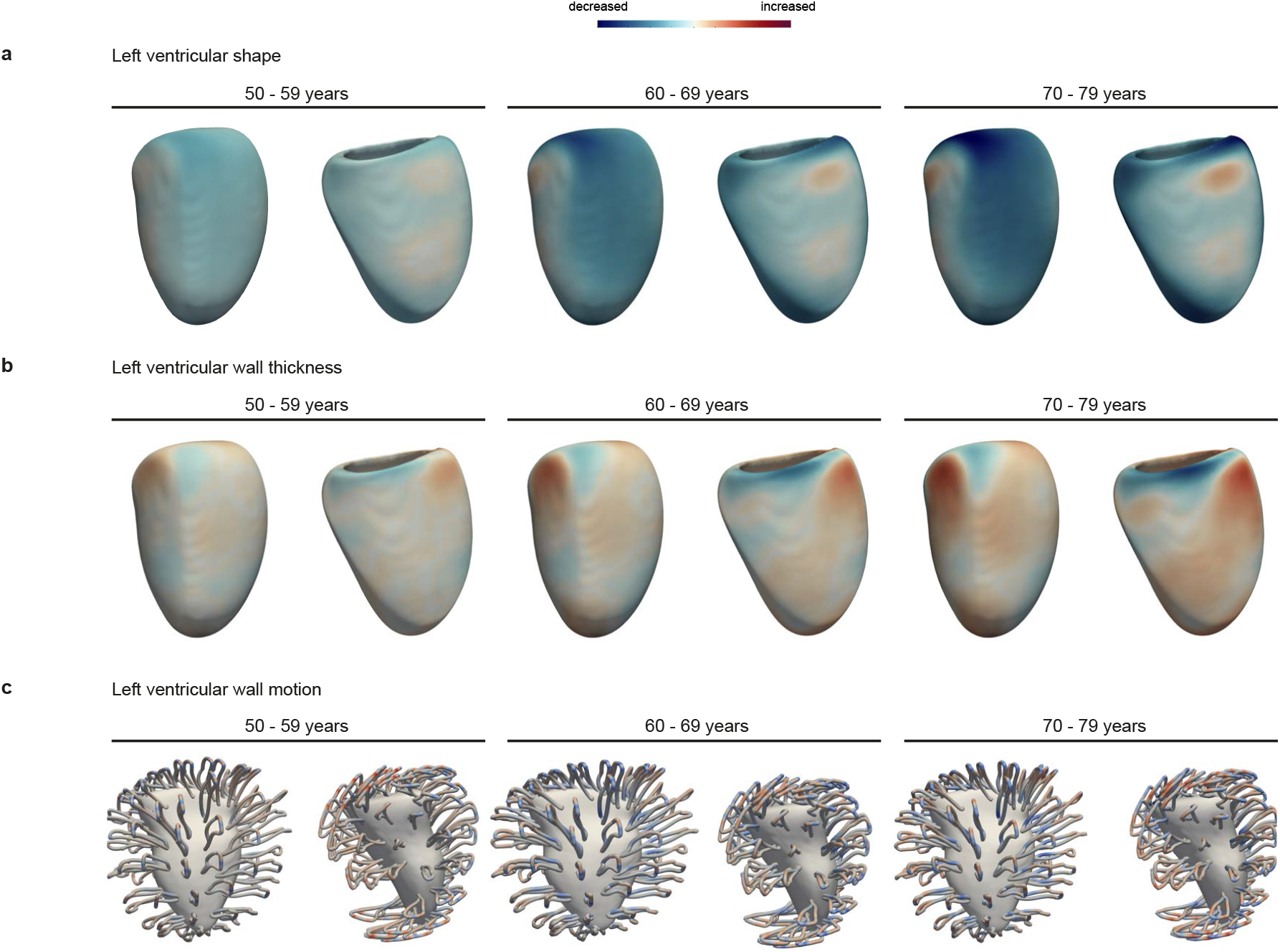
Three dimensional models of cardiac ageing. Three dimensional mapping of left ventricular shape (**a**), left ventricular wall thickness (**b**) and left ventricular motion (**c**) with increasing age. The models show the mean phenotype for each decade of age relative to 40 - 49 year olds, aggregating data using registration of cardiac segmentations, with parameters represented on the epicardial surface. Paired views of the infero-lateral and antero-septal walls.

