## Supplementary Material for "Environmental and genetic predictors of human cardiovascular ageing"

### Supplementary Methods

#### Online methods

The code for each analysis step including image processing, imputation, CatBoost training, risk factor and outcome modelling, and all genetic analyses are presented in Notebook format on GitHub (https://github.com/ImperialCollegeLondon/cardiovascular_ageing).

#### Imputation of data, statistical analyses and performance of predictive models

We used imputation for handling missing values in image-derived phenotypes using multiple imputations with predictive mean matching, with 5 imputations and 5 iterations from the R package `MICE`.[69] For continuous risk factor analysis (i.e. associations of age-delta with alcohol and smoking consumption, and telomere length), we included participants with complete data only. Statistical tests used were two-sided unless otherwise specified.

In Supplementary Table 1, we present an age prediction performance comparison among distinct algorithms evaluated on a holdout set of healthy participants. The CatBoost gradient boosting framework was considered as it natively handles categorical variables, uses ordered boosting to address overfitting, and is rapid to execute at scale on a graphics processing unit (GPU). Here we compare CatBoost, used in the analysis of this paper, with ridge regression, least absolute shrinkage and selection operator (LASSO), elastic net and random forests algorithms. We also report the mean absolute error (MAE), mean squared error (MSE) and coefficient of determination ($R^2$), with their standard errors presented in parenthesis (for the MAE and MSE metrics). The CatBoost model had the best performance metrics. Supplementary Figure 1 demonstrates the prediction using this model on the healthy participant holdout set.

**Supplementary Table 1.** Age prediction performance comparison across algorithms with standard errors presented in parenthesis.

| | MAE | MSE | $R^2$ |
|---|---|---|---|
| CatBoost | 4.207 (0.098) | 27.776 (1.175) | 0.493 |
| Ridge | 4.323 (0.103) | 29.800 (1.321) | 0.456 |
| LASSO | 4.327 (0.101) | 29.432 (1.299) | 0.463 |
| Elastic net | 4.337 (0.101) | 29.501 (1.296) | 0.461 |
| Random forests | 4.614 (0.103) | 32.248 (1.291) | 0.411 |

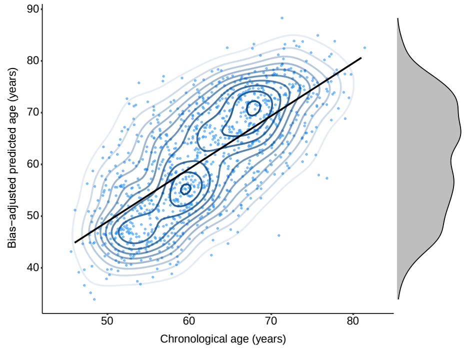

**Supplementary Figure 1. The relationship between predicted and chronological cardiovascular age in the healthy training test set.** Ages jittered, density contours, linear model with 95% confidence intervals, and a marginal density plot, (n=1044).

#### Risk factor and outcome modelling

**Definition of healthy populations, evaluation set population and disease groups in the UK Biobank:** Age is defined as age of participant at the date of the cardiac magnetic resonance (CMR) study, and computed as the number of years between year of birth (UK Biobank data-field 34, https://biobank.ctsu.ox.ac.uk/crystal/field.cgi?id=34) and date of CMR (UK Biobank data-field 53, instance 2, https://biobank.ctsu.ox.ac.uk/showcase/field.cgi?id=53). Sex of participant is determined using UK Biobank data-field 31, (https://biobank.ctsu.ox.ac.uk/crystal/field.cgi?id=31).

Disease definitions were treated as binary traits, and within the UK Biobank were defined based on records from hospital episode statistic (HES) data and self-reported data. If any of the ICD9/ICD10 codes and operation or procedure codes where relevant (for HES data), and any of the self-reported codes (UK Biobank data-field 20002) are reported at any time, we considered the participant to have the disease in question.

In order to define a healthy population relevant to our study of cardiovascular ageing, the UK Biobank participants with CMR were filtered to exclude those with cardiac disease, chronic respiratory disease and metabolic disease (hypercholesterolaemia, diabetes and obesity) using codes defined in Supplementary Table 2, and BMI <30. The remaining participants with CMR were held out to comprise an evaluation set.

Groups of various relevant risk factors were also defined using the codes listed in Supplementary Table 2. For the clinical groups (hypercholesterolaemia, diabetes, obesity, hypertension and coronary artery disease), participants could be part of more than one group. Restricting participants to mutually exclusive disease groups resulted in very small sample sizes without common co-morbidities. Telomere length, smoking and alcohol consumption were obtained using codes documented in Supplementary Table 2. Additionally, alcohol was computed as grams consumed per day[70].

**Definition of health outcomes:** In order to obtain date of clinical health outcome, the "first-occurrence" field (UK Biobank category-field 1712) was used. Time to event was then calculated as number of days between CMR and event. If no event had occurred, participants were censored and time to last data refresh (2021-02-18) used. The primary clinical outcome used was a composite of major adverse cardiovascular events (MACE) and death attributed to cardiovascular cause. Codes used for these definitions are detailed in Supplementary Table 3. Clinical outcomes for 34,133 participants were analysed. Participants had a median time to event or censor of 3.0 years (interquartile range: 2.1-4.4 years) with a total of 732 composite clinical events reported.

**Supplementary Table 2.** A list of diagnostic codes used to define risk factors in the UK Biobank analysis.

| Title | ICD10 Code | ICD9 Code | Self-Reported Code | Operation Code | UK Biobank Field |
|---|---|---|---|---|---|
| **Hypercholesterolaemia** | E78 | 27202, 27209, 27200, 2720 | 1473 | | |
| **Diabetes Mellitus** | E10-E14 | | 2443, 1220, 1221, 1222, 1223, 1276, 1468, 1607 | | |
| **Obesity** | | | | | 21001 |
| **General presence of cardiac disease** | I00-I25, I30-I52 | | | | |
| **Hypertension** | I100, I110, I119, I120, I129, I130, I131, I132, I139, I150, I51, I152, I159 | 4010, 4011, 4019, 4039 | 1065, 1072, 1073 | | |
| **Coronary Artery Disease** | I21-I25 | 4109, 4119 | 1074, 1075 | 1523, 1070, 1071, 1095 | |
| **Chronic respiratory disease** | J40, J410, J411, J418, J42, J431, J432, J438, J439, J440, J441, J448, J449, J450, J451, J458, J459, J46, J47 | 4241 | 1490, 1586 | | |
| **z-adjusted Telomere Length** | | | | | 22192 |
| **Smoking** | | | | | 20161 |
| **Alcohol** | | | | | 4407, 4418, 4429, 4440, 4451, 4462, 1568, 1578, 1588, 1598, 1608, 5364 |

**Supplementary Table 3.** A list of diagnostic codes used to define major adverse cardiovascular event (MACE) outcomes in the UK Biobank analysis.

| Title | ICD10 Code | ICD9 Code | Self-Reported Code | Operation Code | UK Biobank Field |
|---|---|---|---|---|---|
| **Stroke** | I60-I69 | | 6150 | | |
| **Arrhythmia** | I440, I441, I442, I443, I444, I445, I446, I447, I450, I451, I452, I453, I454, I455, I456, I458, I459, I460, I461, I469, I470, I471, I472, I479, I480, I481, I482, I483, I484, I489, I490, I491, I492, I493, I494, I495, I498, I499 | 4260, 4261, 4263, 4264, 4265, 4266, 4267, 4269, 4270, 4271, 4272, 4273, 4274, 4275, 4276, 4278, 4279 | | | |
| **Heart Failure** | I255, I420, I421, I422, I423, I424, I425, I426, I427, I428, I429, I430, I431, I432, I438, I500, I501,I509 | 4251, 4254, 4280,4281,4289 | 1076, 1079 | | |
| **Cardiovascular Death** | Starts with “I” | | | | 40000 |

**Evaluating associations of cardiovascular age-delta with risk factors.** For categorical risk factor groups (i.e. presence or absence of disease), participants with the risk factor ("disease participants") were 1:1 propensity matched with participants without the risk factor ("control participants"), by age and sex. This was performed in R using the package `matchit`.[71] The imbalance of age and sex between both samples was first assessed and quantified using a student's t-test. Next, each disease participant was paired with a control participant with the closest logistic regression propensity score through nearest neighbor matching. The distribution of age and sex was again compared between groups, and additionally propensity scores plotted to visually assess matches qualitatively. Following adjustment for age, age$^2$ and sex, a linear regression model to test for the association between cardiac age-delta and presence of disease as follows:

$$\text{Age delta} \sim \text{Disease (0/1)} + \text{Age} + \text{Age}^2 + \text{Sex} \quad (1)$$

where the coefficient of "Disease" is the effect of having the disease in terms of age delta years.

For continuous risk variables (i.e. alcohol and smoking consumption, and telomere length), we fit a linear regression model in all participants of the test data set that had complete data for these variables to assess the association between cardiovascular age-delta and risk factor as follows:

$$\text{Age delta} \sim \text{Continuous risk factor (e.g. smoking pack years)} + \text{Age} + \text{Age}^2 + \text{Sex} \quad (2)$$

where the coefficient of "continuous risk factor" is the effect of an increase in unit of continuous risk factor in terms of age delta years. No correction for multiple testing was done as we only assessed risk factors with established *a priori* evidence.

**Association of cardiovascular age-delta with outcomes:** We assessed the association of cardiovascular age-delta with prospective cardiovascular events. Non-standard covariates are defined as described in Supplementary Table 2. We first calculated cumulative incidence curves,[72] using death as a competing event for comparing quartiles of cardiovascular age-delta. This is a descriptive analysis which does not include any covariates. We note that no clear split of the curves is visible, indicating that, in an unadjusted analysis, there is no clear association of cardiovascular age-delta with prospective MACE events (see Supplementary Figure 2a).

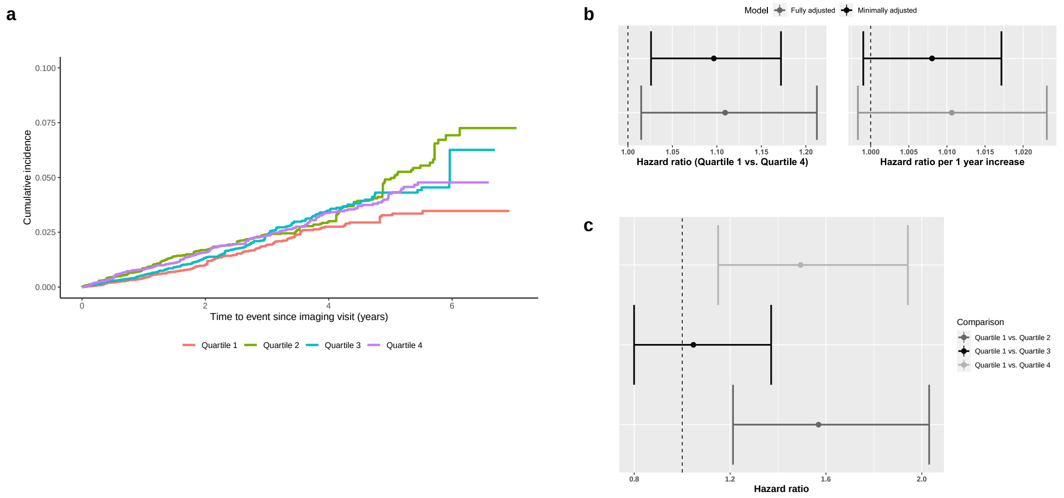

**Supplementary Figure 2. Relationship of cardiovascular age-delta and outcomes. a**, Cumulative incidence curves for time-to-first major adverse cardiovascular event, split into quartile groups based on cardiovascular age-delta. Figure is cut at 97.5 % percentile of reported survival times. **b**, Comparison of first and fourth quartile in a fully adjusted and minimally adjusted model and change in risk per 1 year change in cardiovascular age-delta (data are presented as hazard ratio point-estimates with 95% confidence intervals). **c**, Comparison of quartiles of cardiovascular age-delta. The respective quartile groups are compared to the first quartile which consists of participants with lowest cardiovascular age-delta (data are presented as hazard ratio point-estimates with 95% confidence intervals).

Next, we assessed the association with a Cox model. We consider a fully covariate-adjusted model (age, sex, obesity, diabetes, smoking, alcohol, consumption, hypertension, coronary artery disease, hypercholesterolaemia) and a minimally covariate-adjusted model (age, sex). MACE events prior to the MRI visit are taken into account by a stratified analysis. We compare the lowest and highest quartile (Supplementary Figure 2b) and modelled cardiovascular age-delta as a continuous variable (Supplementary Figure 2c). Although the quartile comparison of the most extreme quartiles yields significant results (hazard ratio 1.0861, $P = 0.01662$), the association does not hold if we model cardiovascular age-delta as a continuous variable.

Considering the cumulative incidence plot (Supplementary Figure 2a), this may be due to a inconsistent pattern across the quartiles. Quartile 2 shows the highest risk for MACE events as well as in the fully covariate-adjusted model (Figure 2d). Therefore, we note that additional analysis are required for strengthening the observed risk profiles of participants with low and high cardiovascular age-delta, and would likely be complemented by longer follow-up data.

**Associations of cardiovascular age-delta with self-reported medication:** In order to assess the potential use of cardiovascular ageing as a surrogate marker of cardiovascular disease progression, we also examined the relationship between self-reported medication and cardiovascular-age delta. We considered the most commonly used drugs, for which effectiveness on clinical outcomes is established: all major antihypertensives, i.e. beta blockers, angiotensin-converting-enzyme inhibitors (ACEi), angiotensin receptor blockers (ARBs), calcium channel blockers (CCBs) and diuretics, as well as statins, metformin and digoxin (Supplementary Table 4).

To assess the association between self-reported medication intake and cardiovascular age-delta, we performed separate drug-specific linear regression models with age-delta as the dependent variable. We adjust for sex, age (linear and quadratic), packs of cigarettes smoked per year, alcohol consumption in grams per day, body-mass index, systolic and diastolic blood pressure (SBP/DBP), heart rate, prior diagnosis of obesity, coronary artery disease, hypertension, diabetes mellitus, hypercholesterolemia and heart failure. All covariates were evaluated at the time of the imaging visit. We conduct a complete case analysis, with respect to the above covariates, which leads to the inclusion of 27,546 participants.

In models adjusting for baseline demographics and key confounding factors, we observed lower cardiovascular age in participants taking CCBs, BBs and ACEis, no significant difference in participants taking statins, diuretics, digoxin, and ARBs, and higher cardiovascular age in participants taking metformin (Supplementary Figure 3). After adjusting estimates for heart rate, SBP and DBP, which are covariates influenced by hypertensive medications to have favourable effects on cardiovascular outcomes, estimates became non-significant with the exception of beta-blockers, where the effect direction changed.

Our models unadjusted for heart rate, SBP and DBP had a $R^2$ of 0.109, which attenuated when models were adjusted ($R^2$ = 0.025). This highlights the contribution of heart rate, SBP and DBP in explaining cardiovascular age-delta.

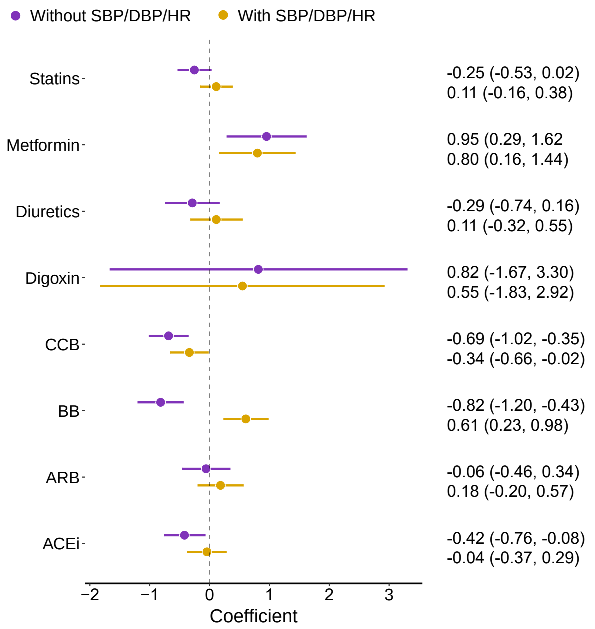

**Supplementary Figure 3. Associations of cardiovascular age-delta with self-reported medication**. Beta-coefficient and 95% confidence intervals are displayed for the association of cardiovascular age-delta and various cardiovascular drugs, adjusted for demographics, lifestyle factors, cardiovascular risk factors and derived using linear regression. Point-estimates are shown for two models: with or without additional adjustment for systolic- and diastolic blood pressure (SBP, DBP) and heart rate (HR). Abbreviations: CCB, calcium channel blockers; BB, beta-blockers; ARB, angiotensin receptor blockers; ACEi, angiotensin converting enzyme inhibitors.

| Characteristic | | | | | $P$ value |
|---|---|---|---|---|---|
| Age-delta quartiles | $[-32.4, -5.3]$ | $[-5.3, 0.2]$ | $[0.2, 5.3]$ | $[5.3, 31.9]$ | |
| N | 6887 | 6886 | 6886 | 6,887 | |
| Diuretics | 252 (3.7%) | 316 (4.6%) | 314 (4.6%) | 348 (5.1%) | <0.001 |
| CCB | 543 (7.9%) | 737 (11%) | 742 (11%) | 752 (11%) | <0.001 |
| ARB | 339 (4.9%) | 383 (5.6%) | 452 (6.6%) | 468 (6.8%) | <0.001 |
| ACEi | 520 (7.6%) | 627 (9.1%) | 599 (8.7%) | 666 (9.7%) | <0.001 |
| BB | 480 (7.0%) | 499 (7.2%) | 491 (7.1%) | 448 (6.5%) | 0.3 |
| Digoxin | 8 (0.1%) | 8 (0.1%) | 9 (0.1%) | 11 (0.2%) | 0.9 |
| Metformin | 189 (2.7%) | 232 (3.4%) | 244 (3.5%) | 302 (4.4%) | <0.001 |
| Statins | 1,498 (22%) | 1,686 (24%) | 1,840 (27%) | 1,670 (24%) | <0.001 |
| Sex | | | | | <0.001 |
| Female | 3,681 (53%) | 3,477 (50%) | 3,395 (49%) | 3,284 (48%) | |
| Male | 3,206 (47%) | 3,409 (50%) | 3,491 (51%) | 3,603 (52%) | |
| Age | 63 (56, 69) | 65 (58, 71) | 66 (60, 71) | 64 (59, 69) | <0.001 |
| Cardiovascular age-delta | -9.0 (-11.9, -7.0) | -2.5 (-3.9, -1.1) | 2.6 (1.4, 3.9) | 8.7 (6.9, 11.3) | <0.001 |

**Supplementary Table 4. Prevalence of self-reported medications across cardiovascular age-delta quartiles**. This table shows the prevalences of self-reported drug use across cardiovascular age-delta quartiles. *N* denotes the number of participants in the corresponding quartile. *P*-values for drugs and sex were determined based on Pearsons Chi-squared test. The *P*-values for age and age-delta were calculated based on Kruskal-Wallis rank sum test. The table is derived on the complete-case set of participants used in the linear regression analyses of medication effects. Abbreviations: CCB, calcium channel blockers; BB, beta-blockers; ARB, angiotensin receptor blockers; ACEi, angiotensin converting enzyme inhibitors.

#### Genotyping and whole exome analysis

**Quality Control of genetic data:** We used imputed genotypes and exome data as described by UK Biobank.[73–75] Participants of European ancestry only have been included and we followed quality control (QC) procedures recommended by UK Biobank, excluding participants as follows:

- Participants who withdrew consent for further analyses were removed (n=5).
- Outliers for heterozygosity or missing rate were removed (UK Biobank field 22027) (n=64).
- Discordance between genetically determined sex and self-reported sex or high missing rate (UK Biobank field 22001 and 31) (n=24).
- Participants with sex chromosome aneuploidy (UK Biobank field 22019) (n=18).
- Close relationships to others, determined by kinship coefficients by UK Biobank > 0.884 (n=2837). One of each pair was excluded at random.

We thus obtained 39,559 participants with CMR data, of which 37,177 participants were of European origin. Applying the above QC filter criteria left 29,506 participants. Finally, these were then divided by date of UK Biobank data release into a discovery set of 20,058 participants (first two releases of imaging data) and a validation set of 9448 participants (third release of imaging data).

We additionally performed a GWAS for ECG-predicted cardiovascular age-delta. Of 43,923 participants with complete ECG 12-lead trace data, 41,122 participants were of European ancestry. Following application of the above QC criteria, 31,475 participants remained. We split the samples analogously to the discovery and validation sets in the CMR-predicted age-delta GWAS.

**Ancestry estimation:** Hidden population substructure is a source of confounding hence we only included participants to comprise a well-mixed population. The majority of participants in the UK Biobank self-identify as white European. Using the R package `plinkQC` (v0.3.3), we used HapMap III reference data for ancestry estimation.[76] HapMapIII reference data was downloaded from ftp://ftp.ncbi.nlm.nih.gov/hapmap/genotypes/2009-01_phaseIII/plink_format/, joined to UK Biobank genotype data and principal component analysis performed. Participants that clustered with HapMap III participants of European ancestry of the PC1/PC2 plot were retained for analyses.

**SNP heritability computation using Linkage Disequilibrium score regression:** To estimate heritability index ($h^2$) we use the Linkage Disequilibrium (LD) score regression tool ("LDSC") obtained from https://github.com/bulik/ldsc. As input we used our GWAS summary statistics for the CMR and ECG GWASs and compute heritability as described in the wiki page https://github.com/bulik/ldsc/wiki/Heritability-and-Genetic-Correlation. We obtain $h^2 = 0.105$ for CMR summary statistics, and $h^2 = 0.1039$ for ECG summary statistics.

**Evidence for variant annotations:** Genome wide significant loci were mapped to estimated causal genes using the following criteria:

- Using VEP annotation, assessment of evidence that the variant directly affects the function of a candidate gene (i.e. predicts damage or loss of function).
- Assessment of whether the variant is an eQTL for the candidate gene in a biologically relevant cell class or tissue. The presence of a colocalising e/pQTL was determined by searching the following resources: GTEx, CEDAR, Fairfax 2012, TWINSUK, eQTLGen, GENCORD, HIPSCI, GEUVADIS, Alasso 2018, Nedelec 2016, Blueprint, Quach 2016, Naranbhai 2015, Van de Bunt 2015, and Schwartzentruber 2018. Locus full summary statistics were downloaded for possible colocalising eQTLs.
- Evidence of additional functional data linking the variant to the gene, using https://genetics.opentargets.org/
- Assessment of the candidate gene's function and plausible contribution to cardiovascular ageing, with further review of Phenome Wide Association Study (PheWAS) associations.

Based on this criteria, the following variant-to-gene mappings were made for the 5 genome wide significant loci (see Supplementary Figure 4 for co-localisation locus plots):

- **rs2042995** eQTL data strongly suggest that the causal gene here is *TTN*. The lead variant, rs2042995, a missense variant found within the *TTN* coding sequence, is also associated with *TTN* expression in blood, atrial appendage, coronary artery, aorta and tibial artery. https://genetics.opentargets.org/variant/2_178693639_T_C

  Summary: Effect variant; increases cardiovascular age delta, probable causal gene: *TTN*.
- **rs7795735** This is an intergenic variant located 12.6 kilobases downstream of *ELN*, the gene that encodes elastin. This was a lead SNP associated with ascending aortic distensibility in a recent publication[77].

  Summary: Effect variant; increases cardiovascular age delta, probable causal gene: *ELN*.
- **rs1991860** This is an intronic variant for *PI15*, which is an eQTL for *PI15* in the tibial artery (GTEx). *PI15* is the closest gene to the locus. *PI15* encodes the peptidase inhibitor 15, which is associated with refractive errors and lung function (https://platform.opentargets.org/target/ENSG00000137558/associations/). There are no clear mechanistic links between the gene and cardiovascular ageing so the conclusion here is inconclusive.

  Summary: Effect variant; decreases cardiovascular age delta. Probable causal gene: *PI15*.
- **rs61886308** This is an intronic variant for *PLCE1*, and is also an eQTL for *PLCE1* in the coronary and tibial artery (GTEx) (Supplementary Figure 4d). *PLCE1* is the closest gene to the locus. Phospholipase C epsilon modulates beta-adrenergic receptor-dependent cardiac contraction and it has been found that this protein is over expressed during heart failure. https://genetics.opentargets.org/variant/10_94146109_G_A

  Summary: Effect variant; increases cardiovascular age delta. Probable causal gene: *PLCE1*.
- **rs2986036** This is an intronic variant for *NEURL1-AS1*, and is also an eQTL for *NEURL1-AS1* in the testes and fibroblasts (GTEx) (Supplementary Figure 4e). *NEURL1-AS1* is the closest gene to the locus. *NEURL1-AS1* was found to be associated with platelet aggregation response to adrenaline.[78] .

  Summary: Effect variant; decreases cardiovascular age delta. Possible causal gene: *NEURL1-AS1*.

**Co-localisation plots:** We performed eQTL co-localisation analysis and show identified eQTLs for our GWAS hits in Supplementary Figure 4.

**Rare variants gene burden analysis:** Supplementary Table 5 shows that significant effect on the age-delta phenotype was found for two genes *TREM2* ($P = 5.01 \times 10^{-6}$, AF<0.1%) and *MICU3* ($P = 7.08 \times 10^{-6}$, on singletons). Leave-one-out-variant (LOVO) test was performed on *TREM2* (Supplementary Table 6). Out of the six LoF variants found in the gene, 6:41158691:C:T (rs2234258), a stop-gained variant, caused the association.

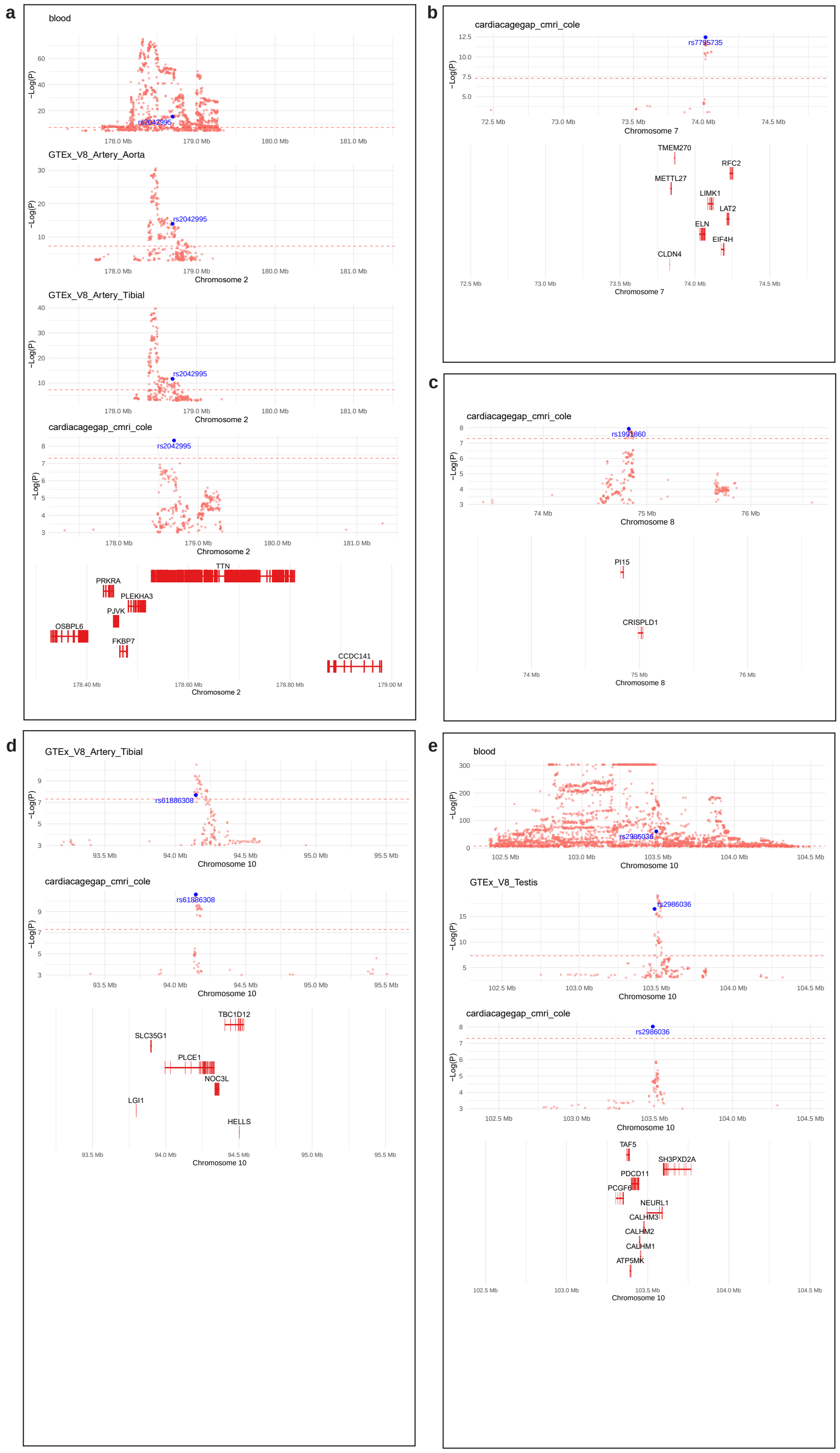

**Supplementary Figure 4. Co-localisation plots of GWAS and eQTL signals at the 5 significant loci.** Co-localisation plots of GWAS and eQTL signals at the 5 significant loci For each of the significant loci, eQTL signals of probable causal genes and some neighboring genes is shown.

#### Polygenic risk score

The polygenic risk score (PRS) was constructed using 29,506 unrelated, European study participants, using SNPs with $P < 0.001$, clumped with LD threshold of $R^2 = 0.1$, window of 250kb. We then constructed a linear (null) model predicting the

**Supplementary Table 5.** Rare variant burden test result on the Age-delta phenotype. Table shows genes with $-log_{10}P > 5$. Masks include M1:LoF, M2:LoF or missense(5/5), M3: LoF or missense(>1/5, 5/5). Analysis was performed using REGENIE on UK Biobank RAP. Number of people tested is 31515.

| CHROM | GENPOS | GENE.MASK.MAF | TEST | BETA | SE | -LOG10P |
|---|---|---|---|---|---|---|
| 6 | 41158607 | *TREM2*(ENSG00000095970).M1.0.001 | ADD | 5.740890 | 1.250900 | 5.35211 |
| 8 | 17027280 | *MICU3*(ENSG00000155970).M1.singleton | ADD | -13.768300 | 3.066220 | 5.14803 |

**Supplementary Table 6.** *TREM2* leave-one-out-variant test.

| CHROM | GENPOS | ID | A1FREQ | BETA | SE | LOG10P |
|---|---|---|---|---|---|---|
| 6 | 41158691 | *TREM2*(ENSG00000095970).M1.0.001_6:41158691:C:T | 0.000079 | 0.337682 | 3.35912 | 0.036247 |
| 6 | 41158994 | *TREM2*(ENSG00000095970).M1.0.001_6:41158994:G:A | 0.000571 | 5.606700 | 1.26863 | 5.004650 |
| 6 | 41158639 | *TREM2*(ENSG00000095970).M1.0.001_6:41158639:GT:G | 0.000571 | 5.715030 | 1.26863 | 5.177760 |
| 6 | 41159883 | *TREM2*(ENSG00000095970).M1.0.001_6:41159883:C:T | 0.000571 | 5.747710 | 1.26861 | 5.230760 |
| 6 | 41158607 | *TREM2*(ENSG00000095970).M1.0.001 | 0.000587 | 5.740890 | 1.25090 | 5.352110 |
| 6 | 41158662 | *TREM2*(ENSG00000095970).M1.0.001_6:41158662:G:A | 0.000571 | 6.156280 | 1.26862 | 5.914430 |
| 6 | 41161557 | *TREM2*(ENSG00000095970).M1.0.001_6:41161557:G:A | 0.000571 | 6.250330 | 1.26869 | 6.077400 |

age-delta using the sex, age (at time of MRI), age$^2$, MRI assessment centre and genotyping array as covariates. $R^2$ of the null model was 0.01. This is compared to a model that adds PRS as an additional covariate. The fraction of variance explained by PRS (Supplementary Table 7) is reported as $\Delta R^2$, and equates to the difference of $R^2$ between these two models.

**Supplementary Table 7.** Results of polygenic risk score (PRS) construction for cardiovascular age-delta using various single nucleotide polymorphism inclusion significance thresholds, with corresponding fraction of variance explained by PRS $\Delta R^2$.

| *P*-value SNP inclusion threshold | Number of SNPs used | $\Delta R^2$ |
|---|---|---|
| $1 \times 10^{-6}$ | 13 | 0.011422831 |
| $1 \times 10^{-5}$ | 45 | 0.029006171 |
| $1 \times 10^{-4}$ | 284 | 0.112262807 |
| $1 \times 10^{-3}$ | 1845 | 0.351992651 |
| $1 \times 10^{-2}$ | 13592 | 0.658321675 |

#### PRS age-delta PheWAS results

In 373,948 genotyped European ancestry UK Biobank participants the age-delta PRS was positively associated with hypertension (OR 1.01, $P = 2.9 \times 10^{-12}$). Coeliac disease was also a significant negative association (Supplementary Figure 5).

#### Age prediction using resting electrocardiograms

**Model specification:** We used a convolutional residual neural network architecture,[79] to predict the age of UK Biobank participants, given as input the corresponding resting electrocardiogram (ECG) traces. The model code and weights were obtained from https://github.com/antonior92/ecg-age-prediction.

**Data preparation:** UK Biobank resting ECG traces were parsed from the XML files provided on the Research Analysis Platform. In these data, one sample corresponds to a measured trace for 1 second at 500 Hz. We downsampled to 400 Hz by uniformly selecting 400 data points for each sample. Afterwards, we selected 4096 data points starting from sample 2. Input data were scaled to be on 1e-4 V scale by dividing the input traces by 10. This preprocessing meets the required input format for the algorithm.

**Training the model:** To adapt the prediction model to the UK Biobank traces, we performed a series of fine-tuning runs. We used our defined healthy participants (see Methods) for training. Performance is evaluated on a held-out validation set of 1000 participants in each training run (roughly 20 percent of the samples). Supplementary Table 8 shows the model-parameters that were used. We present the training results across six different training settings, starting with the pretrained model weights without refinement, full retraining with random and pretrained weights, and fine-tuning of 3 million, 1.7 million and 5120 parameters from the last layers in the network architecture. The models' prediction performances, assessed as Pearson

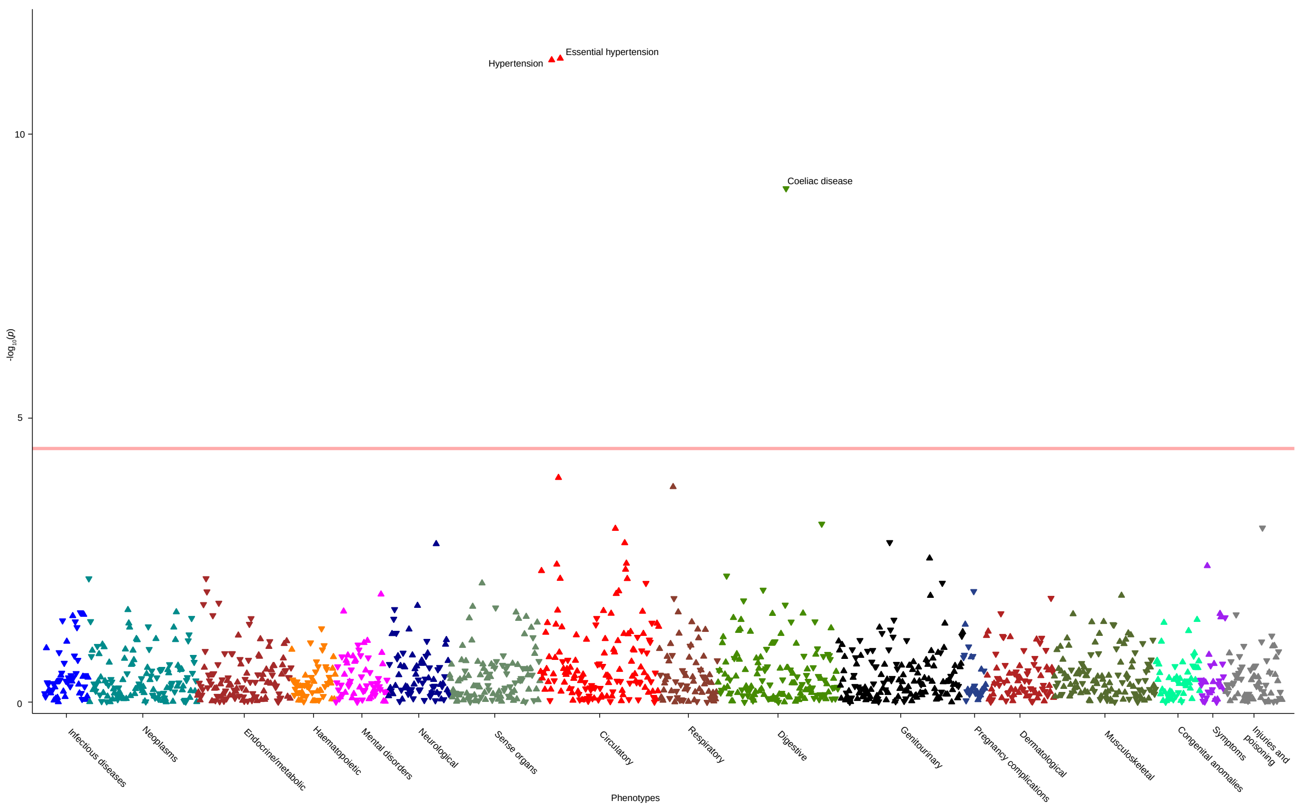

**Supplementary Figure 5. PheWAS using PRS of cardiovascular age-delta** Phenome wide analysis of age-delta PRS adjusted for age, age$^2$, sex and the first ten genetic principal components by logistic regression ($n = 373{,}948$ genotyped White British UK Biobank participants). The red line represents the significance threshold after accounting for multiple testing. Upright triangles indicate positive correlations, and the inverted triangles indicate negative correlations.

correlation coefficient ($|r|$) between predicted age and chronological age at the imaging visit are shown in Supplementary Table 9. Training and validation loss of our selected model **model_pretrained_v3** are shown in Supplementary Figure 6.

| | epochs | random_seed | sample_freq | seq_length | batch_size | lr | patience | min_lr | lr_factor | net_filter_size | net_seq_lengh | dropout_rate | kernel_size |
|---|---|---|---|---|---|---|---|---|---|---|---|---|---|
| 1 | 50 | 112233 | 400 | 4096 | 32 | 0.001 | 7 | 1e-07 | 0.1 | 64,128,196,256,320 | 4096,1024,256,64,16 | 0.8 | 17 |

**Supplementary Table 8. Model parameters for ECG model fine-tuning.** For the training procedure we used these model parameters and left all other parameters as set by default in the ECG model program.

**Prediction performance of ECG age-gap predictions in healthy participants:** As described above, we train our model using 4501 participants, from which we defined a held-out validation set of 1000 random participants serving as performance indicator. We assess the model's performance by the $|r|$ of the predicted vs. chronological age in these 1000 validation participants. Supplementary Figure 7 shows the scatter plots with the predictions. We observe similar prediction performance as the image-based CatBoost model ($|r| = 0.5$, Supplementary Table 9) before age bias correction, and a strong dependency of the age deltas on age, as reported previously. After correction for age-bias,[80] Pearson correlation of chronological age vs. predicted age increases to $|r| = 0.852$ in the healthy validation participants, and age dependency disappears ($r = -0.007$). This is very similar to the results of the image-based CatBoost age delta predictions.

**Prediction of cardiac age-delta from ECGs in UK Biobank:** Given our refined model **model_pretrained_v3** we predict on the remaining 34,108 UK Biobank participants, which, by our definition, do not belong to the "healthy" group. For comparison to image-based predictions, we show the intersection of the 34,108 ECG-based age predictions versus the 34,147 image-based age predictions, leaving in total 28,734 participants. We observe a correlation in the predicted ages (Pearson $|r| = 0.644$), while there is no correlation in age-deltas ($|r| = 0.006$). Supplementary Figure 8 shows the results as scatterplots.

| ModelId | Description | r_NoCorrection | r_ColeCorrected | AgeRange_NoCorrection | AgeRange_ColeCorrected | AgeRange | mean_pred_age | mean_age | mean_delta |
|---|---|---|---|---|---|---|---|---|---|
| model_notrain | Lima et.al. original model | 0.31 | 0.49 | -23.2-88.1 | 2.8-121.4 | 46-81 | 34.60 | 61.80 | -27.20 |
| model_fulltrain_v3 | Full retrain on 5K, random weights | 0.07 | 0.84 | 50-73.9 | 37.4-86.7 | 46-81 | 63.90 | 61.80 | 2.10 |
| model_fulltrain_v2 | Full retrain on 5K, Lima et.al. weights init | 0.54 | 0.80 | 40.6-86.6 | 34-93.3 | 46-81 | 62.50 | 61.80 | 0.70 |
| model_pretrained_v5 | Fine tune 3 Mio parameters from Lima | 0.50 | 0.83 | 40.6-81.3 | 29.4-90.6 | 46-81 | 63.40 | 61.80 | 1.60 |
| model_pretrained_v3 | Fine tune 1.7 Mio parameters from Lima | 0.50 | 0.83 | 40.6-81.2 | 29.4-90.6 | 46-81 | 63.40 | 61.80 | 1.60 |
| model_pretrained_v4 | Fine tune 5120 parameters from Lima | 0.50 | 0.62 | 31.4-89.2 | 26.4-92.7 | 46-81 | 62.40 | 61.80 | 0.60 |

**Supplementary Table 9. Fine-tuning results for ECG training.** The model architecture was retrained in different ways on healthy UK Biobank participants. Above we defined 5066 participant as a 'healthy' population, from which ECG traces from 4501 participants were available. Performance was assessed by correlating the chronological age of the participants at the imaging visit to the predicted age from the model. We perform Cole's method of age-bias correction as described for image-based age prediction described above.[80] Columns presented in this table are: *r_NoCorrection* - Pearson correlation of raw model age predictions to chronological age of participants at imaging visit. *r_ColeCorrected* - Pearson correlation of age-bias corrected age predictions to chronological age. *AgeRange_NoCorrection* - range of predicted ages, no age bias correction. *AgeRange_ColeCorrected* - range of predicted ages with age-bias correction. *AgeRange* - Range of chronological ages at imaging visit. *mean_pred_age* - Mean of predicated ages, *mean_delta* - Mean of predicted 'age-delta', i.e. Predicted age minus chronological Age. We selected a model **model_pretrained_v3** to perform predictions in the rest of UK Biobank.

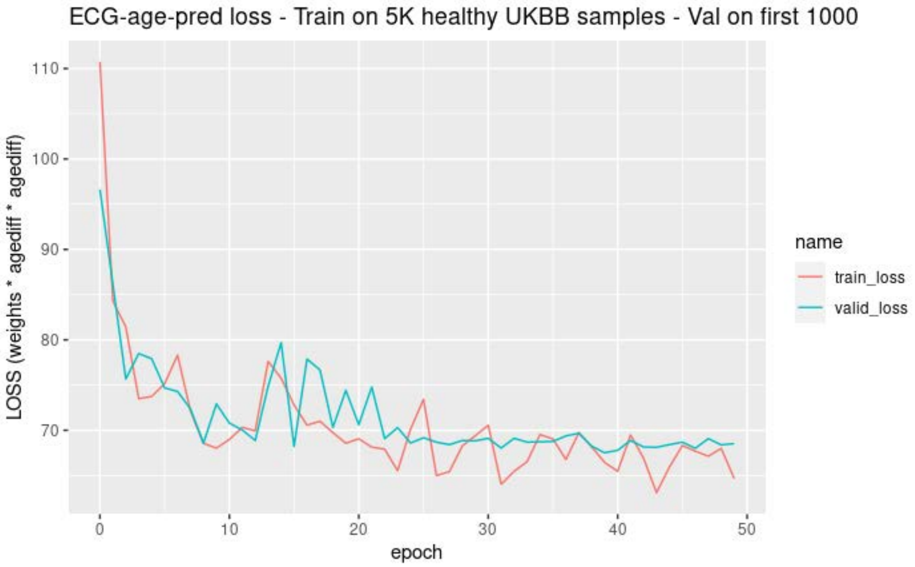

**Supplementary Figure 6.** Training and validation loss of the fine-tuning of our selected ECG model **model_pretrained_v3**.

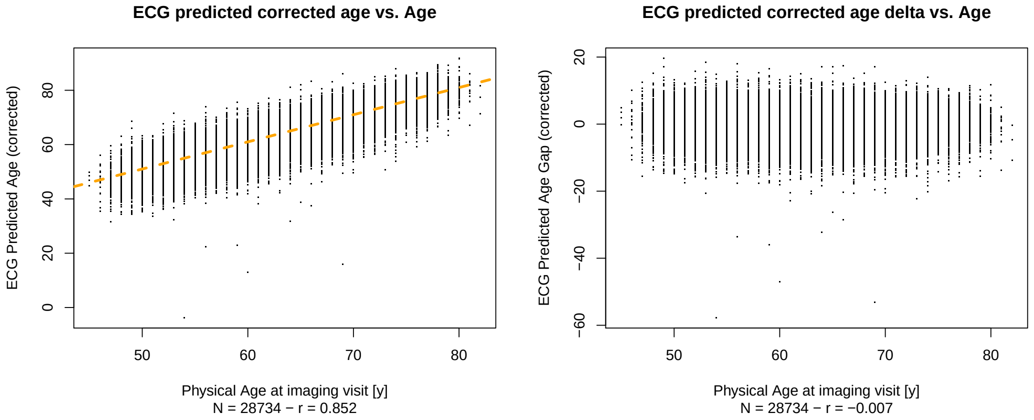

**Supplementary Figure 7.** Training performance assessed as Pearson correlation of predicted vs. chronological age in 1000 healthy held-out participants, using our selected ECG model **model_pretrained_v3**.

**GWAS for ECG predicted age-delta:** Analogous to the GWAS for the CMR predicted age-delta we ran a genome wide association study for the ECG age-delta prediction using PLINK workflow on DNANexus (https://platform.dnanexus.com/app/

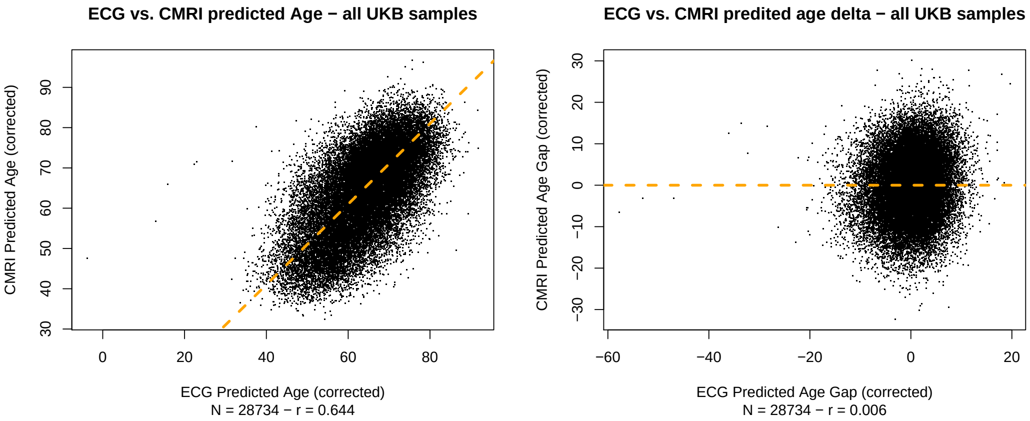

**Supplementary Figure 8.** Comparing prediction of 28734 predicted UK Biobank participants using cardiac magnetic resonance imaging and electrocardiogram (ECG)-based prediction models.

plink_gwas, v1.0.6). We used imputed genotypes in the Bulk/Imputation/UKBiobankimputationfromgenotype folder with the following options:

```
--covar-name Sex,Age,Array,PC1-PC10
--covar-variance-standardize
--geno 0.03
--glm hide-covar
--maf 0.01
--mind 0.05
```

Outputs were collected for each chromosome and a Manhattan plot was generated using a custom ggplot with R. The Manhattan plot is shown in Supplementary Figure 9 and the associated loci in Supplementary Table 10.

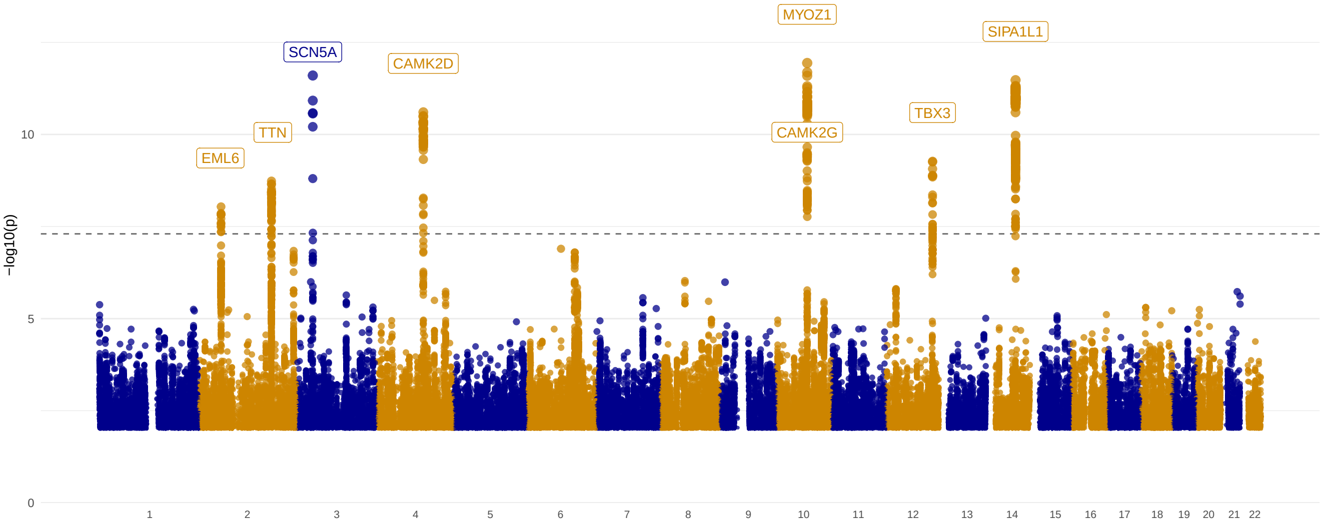

**Supplementary Figure 9.** Manhattan plot for GWAS for ECG based cardiac age-delta prediction. Significance line is at $P \leq 5^{-8}$.

#### Variant annotations:

- **rs4671961** (*EML6*) Summary: Closest gene is *SPTBN1*, but variant is an eQTL for further apart gene *EML6* in tibial, coronary and artery aorta. (https://genetics.opentargets.org/variant/2_54571459_G_A)

- **rs11902709** (*TTN*) Intron variant for *TTN* (closest gene). Identified in both ECG and CMRI based age-delta prediction GWASes. pQTL for proteins FKBP7. PheWAS links to pulse rate and AV-block. (https://genetics.opentargets.org/variant/2_178743480_C_T)

- **rs7373065** (*SCN5A*) *SCN5A* is the closest gene. Other close locus genes are e.g. *SCN10A* and *EXOG*. PheWAS links variant to atrial fibrillation, pulse rate, cardiac arrhythmias. (https://genetics.opentargets.org/variant/3_38668824_T_C)

- **rs35430511** (*CAMK2D*) Closest gene is *CAMK2D*, promoter link in PCHi-C.[81] PheWAS links to atrial fibrillation. (https://genetics.opentargets.org/variant/4_113465982_T_C)

- **rs147790633** (*MYOZ1*) Closest gene is *AGAP5*. eQTL in heart atrial appendage for *MYOZ1*. PheWAS links to atrial fibrillation, cardiac arrhythmias, forced expiratory volume. (https://genetics.opentargets.org/variant/10_73687824_T_C)

- **rs60820984** (*CAMK2G*) Closest gene is *CAMK2G*. Close to rs147790633, and also shows eQTL for *MYOZ1* in heart atrial appendage as this SNP. Linked to promoter in PCHi-C,[81] and DNAse hypersensitive sites.[82] Further linked as enhancer for *NDST2* (*FANTOM5*). PheWAS links to atrial fibrillation, cardiac arrhythmias, peek expiratory flow and lung function. (https://genetics.opentargets.org/variant/10_73879820_C_T)

- **rs7132327** (*TBX3*) Intergenic variant, closest gene is *TBX3* at around 260kb distance. PheWAS links to myocardial fractal dimension. (https://genetics.opentargets.org/variant/12_114943266_T_C)

- **rs35866366** (*SIPA1L1*) eQTL in immune cells and blood and placenta, closest gene *SIPA1L1*. Link to promoter in PCHi-C.[81] PheWAS links to benign neoplasm of other parts of digestive system and myocardial fractal dimension (https://genetics.opentargets.org/variant/14_71382468_A_G)

| **Genome-Wide Association Results for ECG delta** | | | | | | | | | | | | | | | | | |
|---|---|---|---|---|---|---|---|---|---|---|---|---|---|---|---|---|---|
| **Lead variant** | | | | | **GWAS** | | | | | | **Annotation** | | | **Evidence** | | | |
| **rsIDFull** | **Chr** | **Ref** | **Alt** | **MAF** | **Estimate$^{Full}$** | **SE$^{Full}$** | **$P^{Full}$** | **Disc** | **Repl** | **Full** | **Locus genes** | **Closest gene** | **Likely causal gene** | **MS** | **p/eQTL** | **M** | **Overall** |
| rs4671961 | 2 | G | A | 0.3161 | 2.22e-01 | 3.87e-02 | 9.15e-09 | N | N | Y | SPTBN1;EML6 | SPTBN1 | EML6 | | Y | N | Medium |
| rs11902709 | 2 | C | T | 0.0484 | 5.05e-01 | 8.40e-02 | 1.84e-09 | N | N | Y | TTN;PLEKHA3;FKBP7 | TTN | TTN | | Y | Y | High |
| rs7373065 | 3 | T | C | 0.0164 | -9.26e-01 | 1.37e-01 | 1.20e-11 | N | N | Y | SCN10A;SCN5A;EXOG | SCN5A | SCN5A | | N | Y | High |
| rs35430511 | 4 | T | C | 0.2534 | 2.74e-01 | 4.11e-02 | 2.48e-11 | N | N | Y | CAMK2D | CAMK2D | CAMK2D | | N | Y | High |
| rs147790633 | 10 | T | C | 0.1410 | -3.69e-01 | 5.19e-02 | 1.15e-12 | N | N | Y | AGAP5;BMS1P4-AGAP5;MYOZ1 | AGAP5 | MYOZ1 | | Y | Y | High |
| rs60820984 | 10 | C | T | 0.1854 | -2.79e-01 | 4.63e-02 | 1.83e-09 | N | N | Y | CAMK2G;PLAU;NDST2 | CAMK2G | CAMK2G | | Y | N | Low |
| rs7132327 | 12 | T | C | 0.2700 | 2.51e-01 | 4.05e-02 | 5.37e-10 | Y | N | Y | TBX3 | TBX3 | TBX3 | | N | N | Low |
| rs35866366 | 14 | A | G | 0.2435 | 2.88e-01 | 4.14e-02 | 3.35e-12 | N | N | Y | SNORD56B;SIPA1L1 | SNORD56B | SIPA1L1 | | Y | N | Low |

**Supplementary Table 10. Summary of genome wide association study (GWAS) loci from ECG predictions**. For each significant locus, lead single nucleotide polymorphism (SNP) variant information provided (Chr, chromosome; ref, reference allele; Alt, alternative allele; MAF, minor allele frequency), with GWAS summary statistics (Estimate, beta coefficient; SE, standard error; P, P-value). Details of which set the lead SNP reached genome-wide significance presented (Disc, discovery set; Repl, replication set; Full, full dataset). Variant to gene annotation is provided and evidence column summarises MS (missense variant); eQTL (co-localisation of GWAS signal with an expression quantitative trait loci for the gene in a plausible tissue type); M (plausible mechanistic link between the gene and the phenotype i.e. ageing); and Overall (level of confidence of variant to gene mapping given the available evidence).
